# A Transdiagnostic Data-driven Study of Children’s Behaviour and the Functional Connectome

**DOI:** 10.1101/2021.09.15.21262637

**Authors:** J. S. Jones, the CALM Team, D. E. Astle

## Abstract

Behavioural difficulties are seen as hallmarks of many neurodevelopmental conditions. Differences in functional brain organisation have been observed in these conditions, but little is known about how they are related to a child’s profile of behavioural difficulties. We investigated whether behavioural difficulties are associated with how the brain is functionally organised in an intentionally heterogeneous and transdiagnostic sample of 957 children aged 5-15. We used consensus community detection to derive data-driven profiles of behavioural difficulties and constructed functional connectomes from a subset of 238 children with resting-state functional Magnetic Resonance Imaging (fMRI) data. We identified three distinct profiles of behaviour that were characterised by principal difficulties with hot executive function, cool executive function, and learning. Global organisation of the functional connectome did not differ between the groups, but multivariate patterns of connectivity at the level of Intrinsic Connectivity Networks (ICNs), nodes, and hubs significantly predicted group membership in held-out data. Fronto-parietal connector hubs were under-connected in all groups relative to a comparison sample, and children with hot vs cool executive function difficulties were distinguished by connectivity in ICNs associated with cognitive control, emotion processing, and social cognition. This demonstrates both general and specific neurodevelopmental risk factors in the functional connectome.

## 1. Introduction

Behavioural difficulties are common and highly heterogeneous in children who struggle at school (Bathelt, Holmes, et al., 2018). These children may be diagnosed with a neurodevelopmental condition such as Attention Deficit Hyperactivity Disorder (ADHD), autism, and/or dyslexia, but many children have no formal diagnosis (J. Holmes et al., 2019). Whilst behavioural difficulties are key characteristics of multiple different disorder categories they are incredibly heterogeneous in both range and severity (Masi et al., 2017; Wåhlstedt et al., 2009; Willcutt & Pennington, 2000). Even within supposedly singular diagnoses, behavioural profiles can differ markedly (e.g. Bathelt, Holmes, et al., 2018), and some characteristics are apparent across multiple distinct categories (e.g. Kushki et al., 2019).

One good example of this heterogeneity is behaviours related to executive function. Executive functions encompass a broad range of processes that are important for regulating cognition, emotion, and behaviour (Best et al., 2011; Zelazo & Carlson, 2012). Difficulties with executive functions are a common characteristic in multiple neurodevelopmental diagnoses and children who struggle at school (Booth et al., 2010; Demetriou et al., 2018; Frazier et al., 2004; Gathercole et al., 2016; Martinussen et al., 2005). Executive difficulties may manifest as ‘cool’ behaviours associated with poor cognition, ‘hot’ behaviours that are emotionally or motivationally salient (Zelazo & Carlson, 2012), motor behaviours like hyperactivity, social problems with peer relations (C. J. Holmes et al., 2016), and/or learning problems (McClelland et al., 2006). Whilst cool behaviours are considered core to some diagnoses (e.g. ADHD) and hot behaviours to others (e.g. conduct disorder), in reality co-occurrence is common (Hobson et al., 2011; Petrovic & Castellanos, 2016), and they are often apparent in children with no diagnosis at all (Hawkins et al., 2016).

This challenge of heterogeneity has led many to propose an alternative *transdiagnostic* approach to understanding these kinds of difficulties (Bathelt et al., 2018; J. Holmes, Guy, et al., 2020; Siugzdaite, Bathelt, Holmes, & Astle, 2020). With a broader recruitment approach, designed to better capture the full population of children at neurodevelopmental risk, data driven analyses then explore the underlying structure of the data. One common approach is to employ some form of data-driven clustering (e.g. Bathelt, Holmes, et al., 2018; Kushki et al., 2019). These data-driven approaches put to one side diagnostic status, and instead try to link the characteristics themselves with underlying cognitive and neurobiological mechanisms (Archibald et al., 2013; Coghill & Sonuga-Barke, 2012; Ramus et al., 2013). The aim of the current study is to investigate a) whether distinct data-driven profiles can explain the diversity of behavioural difficulties in childhood, and b) how these profiles are associated with functional brain organisation. We use an intentionally heterogeneous sample of children identified as struggling in the areas of attention, learning, and memory, with broad recruitment from multiple referral routes (J. Holmes et al., 2019).

Network science is one data-driven method for understanding the underlying structure of large-scale heterogeneous datasets. In a behavioural network, nodes represent individuals and the edges that connect them represent correlations in behaviour. Distinct ‘communities’ of highly related children can be discovered using algorithms that maximise ‘modularity’, which is the number and strength of within-community edges compared to chance. In a functional brain network, nodes represent individual brain regions and the edges that connect them represent temporal correlations in regional brain activity, known as functional connectivity. The network organisation of functional connectivity in the resting-brain is modular, comprising of multiple intrinsic connectivity networks (ICNs), and it has small-world properties, maximising efficient communication whilst minimising wiring costs (Bullmore & Sporns, 2009). Small-world networks have high average clustering, which is the proportion a node’s neighbours that are also connected to each other, and a short average path length, which is the number of edges joining node pairs, and is inversely proportional to global efficiency (Bassett & Bullmore, 2017).

Individual differences in behavioural difficulties across children and adolescents are associated with variability in the functional interactions within and between ICNs. To continue with the example of executive function difficulties: A key finding is that better executive function and better regulation of inattention, hyperactivity and impulsivity is associated with greater segregation of the default mode network from ICNs implicated in cognitive control and attention. Specifically, better parent ratings of overall executive function are associated with reduced connectivity between the default mode network and cingulo-opercular network (Abbott et al., 2016; although see Hawkey et al., 2018). Much of the literature has focused on behavioural ratings of characteristic ADHD symptoms, including inattention, hyperactivity and impulsivity, which are strongly related to executive function behaviours (Silverstein et al., 2020). Less severe ADHD symptoms are associated with reduced connectivity between the default mode network with the dorsal attention (H. Lin et al., 2018) and ventral attention networks (Sripada et al., 2014). Furthermore, less severe symptoms of ADHD are associated with greater cross-network interaction of the cingulo-opercular network, a combined measure of segregation from the default mode network and integration with the lateral fronto-parietal network (Cai et al., 2018). Mirroring these findings, atypical patterns of connectivity between the default mode, cingulo-opercular, and fronto-parietal networks have been observed in those with neurodevelopmental and mood conditions (Menon, 2011), suggesting that this altered connectivity may be a transdiagnostic marker of behavioural difficulties.

Whilst integration between the cingulo-opercular and fronto-parietal networks may be altered in many diagnoses (Menon, 2011), the relationship with executive function-related behaviours is unclear. Better ratings of overall executive function and ADHD symptoms have been associated with both reduced anti-correlations (i.e. stronger functional connectivity Hawkey et al., 2018) and weaker functional connectivity between the cingulo-opercular and fronto-parietal networks (Qian et al., 2019). This inconsistency may be due to small sample sizes (*Ns*=58-83) and variability in the spatial topologies of the networks analysed. Executive function-related behaviours have been more consistently linked to connectivity within these networks. Weaker connectivity within the cingulo-opercular network has been associated with better ratings of overall executive function (Abbott et al., 2016; Hawkey et al., 2018) and less severe symptoms of ADHD (Yerys et al., 2019). On the other hand, stronger connectivity within the fronto-parietal network has been associated with less severe symptoms of ADHD (Francx et al., 2015; H. Y. Lin et al., 2015). A large study of 229 children found that stronger connectivity within the fronto-parietal network was associated with less hyperactivity and impulsivity over time (Francx et al., 2015), suggesting that better integration within the fronto-parietal network may support behavioural self-regulation.

Hot and cool aspects of executive function may have distinct associations with connectivity (Posner et al., 2013). For example, better working memory ratings have been associated with greater connectivity between regions of the cingulo-opercular and visual networks (Zhao et al., 2017), while hot executive function behaviours are particularly associated with connectivity in the limbic system (Ho et al., 2015; Hulvershorn et al., 2014; Karalunas et al., 2014; Posner et al., 2014). Impulsivity has been associated with reduced connectivity between the cingulo-opercular / dorsal attention network and premotor regions (Shannon et al., 2011), atypical functional connectivity of the nucleus accumbens reward system (Costa Dias et al., 2015), and competitive interactions between the ventromedial prefrontal cortex connections with the dorsolateral prefrontal cortex and the mesocorticolimbic reward system (Zhai et al., 2015). This suggests that impulsive behaviour may be linked to altered co-activation of brain regions implicated in cognitive control with regions implicated in motor control and reward. In summary, these findings broadly suggest that networks required for cognitive control are implicated in behaviours associated with both hot and cool executive function, but that their connectivity with other brain regions may be domain specific.

Only one study has examined the relationship between global properties of the functional connectome and behavioural difficulties in childhood. Inattentive behaviours in children with and without ADHD were associated with reduced strength, clustering, path length, and local efficiency (Wang et al., 2020). However, it is unclear whether these differences in graph theory metrics are a result of overall differences in functional connectivity. Further work is required to establish whether these findings generalise to other child populations, whilst controlling for mean functional connectivity, and how global organisation of the functional connectome relates to other behavioural domains.

Executive function-related difficulties provide one particularly good example of a transdiagnostic behavioural domain that’s highly relevant for understanding heterogeneity across children at neurodevelopmental risk. These difficulties are captured, at least in part, by multiple different behavioural checklists and there is a growing neuroimaging literature exploring different aspects of executive function difficulty, either within or across diagnostic groups. But there are multiple relevant behavioural domains, including socio-emotional processing. Recently, data-driven subtyping across a range of behaviours has been shown to produce more distinct and homogenous groupings, relative to diagnostic status (Bathelt, Holmes, et al., 2018). Indeed these data-driven behavioural groupings are somewhat independent of diagnostic status, but strongly linked with differences in brain connectivity (Bathelt, Holmes, et al., 2018; Karalunas et al., 2014).

### 1.1. The Present Study

The primary purpose of the current study is to investigate the relationship between behavioural difficulties and functional brain organisation. To do this, we will first identify whether a few distinct profiles can reasonably explain the diversity of behaviour in a heterogeneous sample of 957 struggling learners aged 5-15. This will be achieved using community detection to derive behavioural profiles from the Conners 3 parent-report questionnaire. This questionnaire is widely used in clinical contexts because it covers a range of relevant behavioural domains. It is best known for its role in supporting diagnosis of ADHD, owing to its good coverage of both hot and cool executive function behaviours. Whilst parent-reports have known limitations, they seem to capture important variance not captured by specific tasks (Barkley & Murphy, 2010; Biederman et al., 2008).

We will examine whether the behavioural profiles derived from community detection on the Conners 3 subscales are associated with functional brain organisation. While previous studies have often examined regional connectivity, ICNs, or global connectome organisation in isolation, we will examine associations at each of these levels in the same sample. Partial least squares (PLS) regression, a multivariate dimension reduction technique, will be used to identify components that maximally explain covariance between the behavioural profiles and functional connectomes.

## 2. Method

The aims and methods of this registered report were pre-registered after undergoing initial Stage 1 peer-review. These are stored along with the analysis scripts at: https://osf.io/cvsu2.

### 2.1. Sample Characteristics

A total of 957 children aged 5-15 years (*M*=9.52, *SD*=2.31) were recruited from the Centre for Attention Learning and Memory (CALM; J. Holmes et al., 2019). Assent and parental consent were obtained for all participating children. Children were excluded from the study if they had an uncorrected hearing or visual impairment, pre-existing neurological condition, a known genetic cause for their difficulties, or if they were a non-native English speaker. Children in the struggling learners sample (*n*=799) were referred by educational and health practitioners for having one or more difficulties in attention, memory, language, literacy, and numeracy. The comparison sample (*n*=158) was recruited from the same schools but were not identified as struggling. Demographics and diagnostic status of the two samples are provided in Table 1 and age distributions are provided in Supplementary Figure 1.

**Table 1.** Sample Characteristics.

| | Struggling learners ($n=799$) | Comparison sample ($n=158$) |
| --- | --- | --- |
| Age in years: $M$ ($SD$) | 9.42 (2.29) | 10 (2.33) |
| Boys: $n$ | 550 (68.8%) | 89 (56.3%) |
| Girls: $n$ | 249 (31.2%) | 69 (43.7%) |
| No diagnosis: $n$ | 482 (60.3%) | 155 (98.1%) |
| ADHD: $n$ | 194 (24.3%) | 1 (0.6%) |
| Suspected ADHD: $n$ | 57 (7.1%) | 0 (0%) |
| Autism: $n$ | 56 (7%) | 0 (0%) |
| Dyslexia: $n$ | 47 (5.9%) | 2 (1.3%) |

### 2.2. Measures

#### 2.2.1. Behaviour

The Conners Parent Rating Short Form 3^rd^ Edition is a validated and reliable parent questionnaire of behaviour in childhood (Conners, 2013). Parents or carers rated the frequency of 45 behavioural items over the past month across six scales measuring: inattention, hyperactivity/impulsivity, learning problems, executive function, aggression, and peer relations. Ratings on these scales will be used to construct the behavioural network.

#### 2.2.2. Intelligence

The Matrix Reasoning subtest of the Wechsler Abbreviated Scale of Intelligence (WASI-II; Wechsler, 2011) was used as a measure of fluid intelligence. It includes 30 visuospatial reasoning problems and children must choose which of five answers fits with the sequence or pattern. The Peabody Picture Vocabulary Test (PPVT) is a measure of receptive vocabulary (Dunn & Dunn, 2007) and was used here as a measure of crystallised intelligence. The experimenter read aloud progressively more unfamiliar words and the participant chose the corresponding image from four options. Scores were standardised according to age norms (*M*=100, *SD*=15) and were averaged between the two tests. Intelligence scores were only used to characterise different groups of children identified in the behavioural network.

#### 2.2.3. Academic Attainment

The Word Reading and Numerical Operations subtests of the Wechsler Individual Achievement Test (WIAT-II; Wechsler, 2005) were used as measures of Reading and Maths, respectively. Word Reading primarily requires children to read aloud single words that are progressively more unfamiliar. Earlier items require identifying letters, phonemes, and similar sounding words. Numerical Operations primarily requires children to solve arithmetic problems on paper with progressive difficulty. Earlier items include number identification and counting, and advanced items include algebra. Some children (*n*=68) completed the Maths Fluency subtest of the Woodcock Johnson III Test of Achievement (WJ-III; Woodcock et al., 2001), instead of the Numerical Operations. In this test children are given three minutes to correctly answer as many simple maths calculations as possible. All scores were standardised according to age norms (*M*=100, *SD*=15). Maths scores will be combined across both tests but analyses will be checked for robustness against the larger sample that completed the Numerical Operations (*n*=889). Maths and reading scores will only be used to characterise any groups identified in the behavioural network.

### 2.3. Behavioural Network Construction and Profiling

Behavioural profiles were determined by replicating the consensus community detection procedure in Bathelt and colleagues (2018), which was conducted on an earlier sample of 442 struggling learners from the CALM cohort. First, a behavioural network of the struggling learners sample (*n*=799) was constructed by calculating the Pearson’s correlation across the six scales of the Conners 3 questionnaire for every pair of children. The community Louvain algorithm assigned each child to an individual community and then iteratively divided the network into communities to maximise modularity quality (*Q*), which is the number of within community associations relative to random chance (Blondel et al., 2008). A consensus partition was determined across 100 iterations of the community detection (Lancichinetti & Fortunato, 2012). The degree of separation between the behaviour profiles was quantified by modularity. Previously this procedure showed good separation into three groups (*Q*=0.55), which was reliable in a random half (*Q*=0.6) and quarter of the sample (*Q*=0.61), and created more homogenous behavioural profiles compared to traditional diagnostic categories (Bathelt, Holmes, et al., 2018). All behavioural and functional brain network analyses were run in the Brain Connectivity Toolbox (Rubinov & Sporns, 2010) for Python (https://github.com/aestrivex/bctpy).

To characterise the profiles identified by community detection we compared each pairwise combination of the profiles and the comparison sample (Bonfferoni corrected) on each of the Conners scales. We used nonparametric Mann-Whitney *U-*tests assuming within-group scores are non-normal, as previously shown (Bathelt, Holmes, et al., 2018). We also examined group differences in demographics, diagnosis, intelligence, and learning. Here, t-tests (Bonferroni corrected) were used to test group differences in age, intelligence, maths, and reading, and chi square tests were used to test group differences in the frequencies of males/females and common diagnoses (ADHD, dyslexia, autism, and no diagnosis).

### 2.4. Image Acquisition

Magnetic resonance imaging data were acquired at the MRC Cognition and Brain Sciences Unit, University of Cambridge. All scans were obtained on a Siemens 3T Prisma-fit system (Siemens Healthcare, Erlangen, Germany), using a 32-channel quadrature head coil.

In the resting-state fMRI, 270 T2*-weighted whole-brain echo planar images (EPIs) were acquired over nine minutes (time repetition [TR] = 2s; time echo [TE] = 30ms; flip angle = 78 degrees, 3×3×3mm). The first 4 volumes were discarded to ensure steady state magnetization. Participants were instructed to lie still with their eyes closed and to not fall asleep. For registration of functional images, T1-weighted volume scans were acquired using a whole-brain coverage 3D Magnetization Prepared Rapid Acquisition Gradient Echo (MP-RAGE) sequence acquired using 1-mm isometric image resolution (TR = 2.25s, TE = 2.98ms, flip angle = 9 degrees, 1×1×1mm).

### 2.5. fMRI Pre-processing

Only a subset of children opted to take part in the MRI study. We used all of the available resting-state fMRI data, which was present for 349 children. The data was minimally pre-processed in fMRIPrep version 1.5.0 (Esteban et al., 2019), which implements slice-timing correction, rigid-body realignment, boundary-based co-registration to the structural T1, segmentation, and normalisation to the MNI template. The data were then smoothed by 6mm full-width at half-maximum. Many methods exist to denoise motion and physiological artefacts from resting-state fMRI; however, the effectiveness of these strategies varies depending on the sample (Ciric et al., 2017; Parkes et al., 2018). We evaluated the performance of several denoising strategies (head movement regressors, aCompCor, ICA-AROMA, motion spike regression, white matter [WM] and cerebrospinal fluid [CSF] regression, and global signal regression) on several quality control metrics (edge weight density, motion-functional connectivity correlation, distance-dependence, and functional degrees of freedom lost) using the fmridenoise package in Python (Finc et al., 2019; see Supplementary materials). The most effective confound regression procedure included a band-pass filter between 0.01-0.1Hz, 24 head motion parameters (six rigid body realignment parameters, their squares, their derivatives, and their squared derivatives), 10 aCompCor components from the WM and CSF signal (Behzadi et al., 2007), linear and quadratic trends, and motion spikes (framewise displacement >0.5mm; Power et al., 2012). Simultaneous confound regression was performed in the Nipype (version 1.2.0) implementation of AFNI’s 3dTproject (Cox, 1996). Children were first excluded for high average motion (mean framewise displacement >0.5mm, *n*=93) and then for a large number of motion spikes (>20% spikes, *n*=18), where few temporal degrees of freedom would have remained. The final functional connectome sample includes 238 children (struggling learners *n*=175, comparison *n*=63). Average in-scanner motion was 0.2mm (*SD*=0.09mm).

### 2.6. Functional Connectome Construction

The denoised fMRI data were parcellated according to 400 region resting-state fMRI cortical parcellation (Schaefer et al., 2018) and a 64 region subcortical parcellation derived from structural connectivity data (Fan et al., 2016). Pearson correlations were computed for the regional time-series within each individual generating 464×464 connectivity matrices. We used proportional thresholding to remove spurious false-positive edges. This approach is recommended over absolute thresholding as it controls for the number of edges across individuals, which strongly influences many graph metrics (Van Wijk et al., 2010; Váša et al., 2018). Specifically, individual connectivity matrices were thresholded to retain the top 25% of positive edges at the group level, ensuring that the same edges are retained for comparison across individuals in subsequent analyses, as in Baum et al. (2017). Any negative edge weights that survived the group threshold were set to a small positive value (0.001) to minimise their influence. To test the robustness of any significant brain-behaviour results, connectomes were generated at additional cost thresholds (5%, 10%, 15%, 20% and 30%) and the Area Under the Curve (AUC) was examined. As a further test of robustness, global graph metrics were also computed for individually thresholded connectomes, as these may capture greater individual variability in global brain organisation. Edge weights were normalised to the maximum value within individuals’ connectomes. Outlier analysis was performed for mean functional connectivity and cases were removed if they were three standard deviations away from the mean. Average functional connectivity was calculated within and between eight pre-defined ICNs: visual, somatomotor, dorsal attention, ventral attention, fronto-parietal, default mode, limbic, and subcortical.

### 2.7. Global Connectome Properties

Global graph theory metrics were computed for the weighted thresholded functional connectomes (Rubinov & Sporns, 2011). Strength is a nodal property that defines the weighted sum of all of a node’s direct connections. The global clustering coefficient is the average proportion of a node’s neighbours that are also connected to one another. The characteristic path length is the average shortest path length between every pair of nodes in the network, global efficiency is the average inverse shortest path length in the network, and local efficiency is the average inverse shortest path length in each node’s neighbourhood. Small-worldness was estimated as the global clustering coefficient divided by the characteristic path length (Bassett & Bullmore, 2017). Assortativity is the correlation between the strength of pairs of connected nodes in the network. Finally, modularity is defined as the weighted proportion of connections within modules compared to that expected by chance. Graph metrics were be analysed at the global connectome level. The following graph metrics were normalized according to the average of 100 random networks with the same degree and weight distribution: global clustering coefficient, characteristic path length, global efficiency, local efficiency, small-worldness, and assortativity.

### 2.8. Brain-behaviour Analyses

The association between the behavioural profiles and functional connectomes was tested at three levels: the global connectome, ICNs, and regions. Global graph properties of the connectome were compared between the behavioural profiles in a series of ANCOVAs, including age, sex, motion, and mean functional connectivity as covariates. Significant main effects were followed up with pairwise ANCOVAs between all behavioural profiles and the comparison sample (Bonferroni corrected).

At the ICN level, dummy variables were created for the behavioural groups using one-hot-encoding and ICN variables were created by averaging edge weights within each ICN and between each pair of ICNs. ICN variables were mean-centered and scaled to unit variance. We then used Partial Least Squares (PLS) regression to evaluate the components of ICN variables that best explain group membership across the behavioural profiles and comparison sample, whilst controlling for age, sex, motion, and mean functional connectivity. The model fit was evaluated by using 5-fold cross-validation repeated 10 times with random splits. The root mean square error (RMSE) from the cross-validated models was compared to permuted null models using 1000 randomly shuffled samples. The contribution of ICN variables to the PLS components was then evaluated using a bootstrap procedure by sampling the total sample size with replacement 1000 times. The loadings onto PLS components was calculated as the mean loading divided by the standard error (SEM) across permutations, where a Procrustes rotation was applied to align the factors across iterations (Krishnan et al., 2011). PLS will be run using scikit-learn 0.22.2 in Python 3.7.3.

To examine how individual brain regions explain the behavioural profiles, we repeated the PLS procedure for node strength. Furthermore, as inter-network segregation and integration can develop simultaneously in hubs (Baum et al., 2017), we examined the unique contributions of hub regions to executive function-related behaviours. Connector hubs were defined as nodes with high betweenness centrality (above the 70th percentile; Baum et al., 2017), which measures how often a node participates in the shortest path between pairs of nodes in the connectome, and high participation coefficient, which measures the diversity of a node’s connection between networks (above the 70th percentile; Power et al., 2013). Provincial hubs were defined as having a high within-module degree (above the 70^th^ percentile) and low participation coefficient (below the 70th percentile; Xu et al., 2016). Here, the PLS procedure was repeated for connector and provincial hubs separately.

## 3. Results

### 3.1. Community Detection

We created a behavioural network from child-by-child correlations across the six scales of the Conners for 777 struggling learners. Twenty-two children were excluded because their scores on the scales did not vary, thereby precluding calculation of the correlation coefficient. Consensus community detection of the behavioural network identified three communities of children, but the composition of these communities varied slightly with repeated iterations of the algorithm. We, therefore, re-ran the algorithm 100 times after removing data from 74 individuals with above-threshold scores on the Negative Impressions scale. This scale indicates an overly negative bias in the parent or guardian’s ratings and may affect the resulting correlations in the behavioural network (Conners, 2013). The resulting communities identified were consistent across all 100 iterations of the algorithm and only six participants had variable community assignments. We then compared the community assignment from this slightly reduced sample with the original results of community detection on the full sample. There was 96.6% agreement with one of the original community assignments from the full sample and the correlations between mean scores across these community assignments exceeded 0.99. Therefore, we were confident using the original community assignments for the whole sample as they aligned very well with the assignments when individuals with negative impressions were removed. These community assignments are shown in Figure 1 and indicated good separation between the groups (*Q*=0.46).

**Figure 1.**
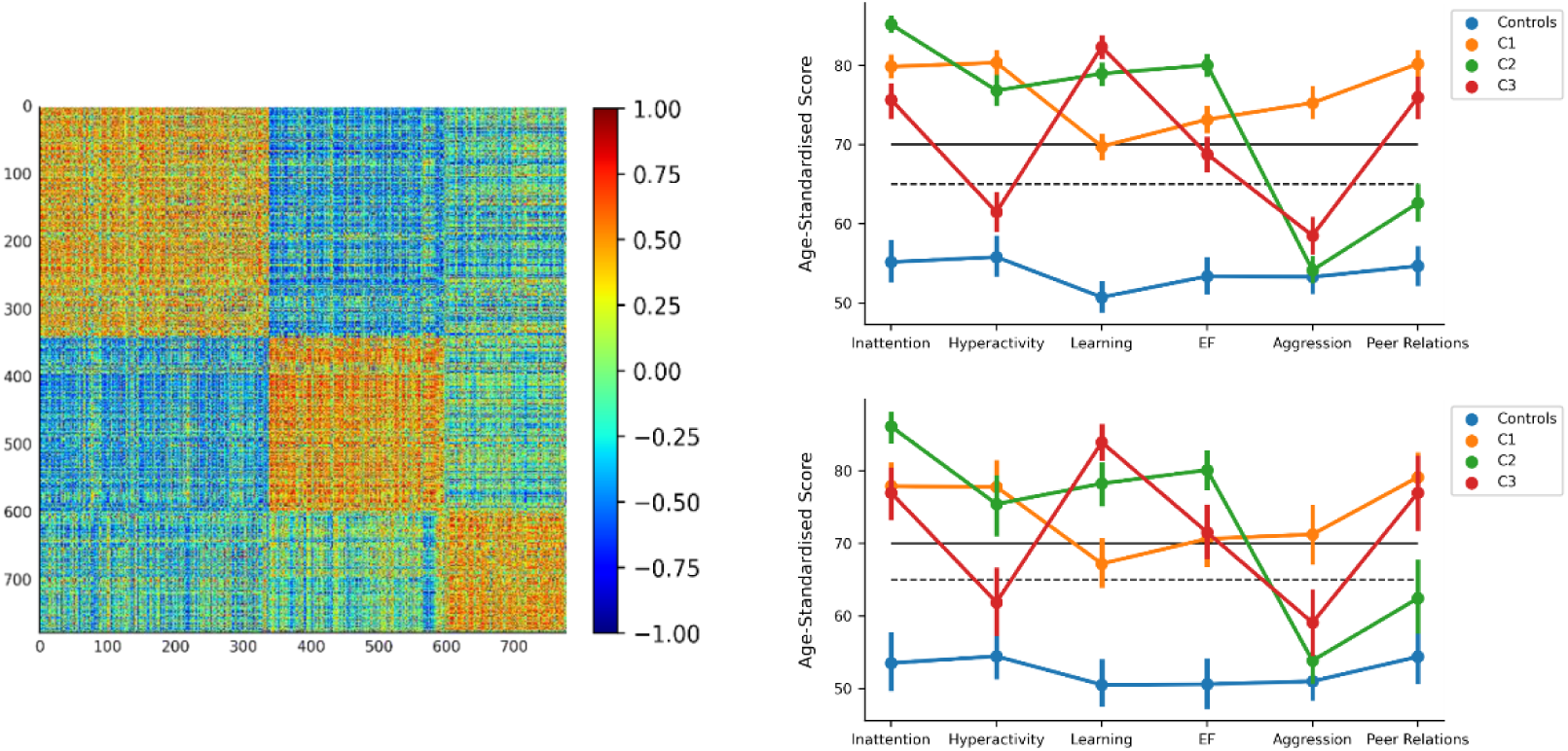
Results of the consensus community detection. The behavioural network (left) shows the child-by-child correlation matrix across the six scales of the Conners 3 after sorting by the community assignments: C1 (*n*=339), C2 (*n*=260), and C3 (*n*=178). The behavioural profiles for the three communities and Controls in the whole sample (top right) and MRI sample (bottom right). Points indicate mean scores and error bars denote 95% confidence intervals from 1000 bootstrapped samples. The dashed horizontal line indicates the threshold for elevated scores and the solid line indicates very elevated scores. Note: Executive Function (EF), Hyperactivity/Impulsivity (Hyperactivity), Learning Problems (Learning).

### 3.2. Behavioural Profiles

The groups showed distinct profiles of behaviour (see Figure 1 & Tables S2-3). C1 (*n*=339) had the highest ratings of Aggression, Peer Problems and Hyperactivity/Impulsivity; C2 (*n*=260) had the highest ratings of Inattention and Executive Function difficulties; and C3 (*n*=178) had the highest ratings of Learning Problems. Mann-Whitney *U-*tests indicated that all pairwise comparisons were significantly different for all behaviour scales (*p* < 0.05 Bonferroni corrected), except for the comparison of C2 and Controls on Aggression. In the MRI sample, these profiles were consistent (see Figure 1 & Tables S4-5), however, some pairwise comparisons were no longer statistically significant (*p* > 0.05 Bonferroni corrected): C1 and C3 on Inattention, C1 and C2 on Hyperactivity/Impulsivity, C1 and C3 on Executive Function, C2 and C3 on Aggression, Controls and C2 on Peer Relations, and C1 and C3 on Peer Relations.

The groups also significantly differed on some demographic, diagnostic, cognitive, and learning outcomes (see Tables 2, S6 & S7). As expected, the control group had significantly fewer diagnoses overall and ADHD diagnoses compared to each data-driven group. There were also significantly fewer diagnoses of autism in the control group relative to C1 and C2, and significantly fewer diagnoses of dyslexia relative to C3. C1 included significantly more children with diagnoses than expected compared to C2, significantly more children diagnosed with ADHD than C2 and C3, and significantly more children diagnosed with autism relative to C2. In contrast, C1 included fewer children diagnosed with dyslexia relative to C2 and C3. C1 also included a significantly greater proportion of boys than all other groups. On measures of cognition and learning, the comparison sample scored higher on IQ, reading, and maths compared to each data-driven group. Of the data-driven groups C1 had the highest scores, which were significantly higher on IQ, maths, and reading than C2 and C3. C3 had the lowest scores, which were significantly lower on IQ and reading than C2. In short, C1 members were more likely to be boys and have ADHD or autism diagnoses. In addition, C1 had the highest cognitive and educational performance of the data-driven groups followed by C2 and then C3.

**Table 2.** Group Characteristics.

|  | Controls | C1 | C2 | C3 |
| --- | --- | --- | --- | --- |
| *N* | 158 | 339 | 260 | 178 |
| Boys: *n* (%) | 89 (56.33%) | 271 (79.94%) | 155 (59.62%) | 110 (61.8%) |
| Girls: *n* (%) | 69 (43.67%) | 68 (20.06%) | 105 (40.38%) | 68 (38.2%) |
| Diagnosis: *n* (%) | 6 (3.8%) | 163 (48.08%) | 83 (31.92%) | 66 (37.08%) |
| ADHD: *n* (%) | 1 (0.63%) | 123 (36.28%) | 46 (17.69%) | 21 (11.8%) |
| Dyslexia: *n* (%) | 2 (1.27%) | 7 (2.06%) | 20 (7.69%) | 19 (10.67%) |
| Autism: *n* (%) | 0 (0%) | 36 (10.62%) | 8 (3.08%) | 13 (7.3%) |
| Age: *M* (*SD*) | 10.85 (2.21) | 10.43 (2.38) | 10.74 (2.09) | 9.82 (2.06) |
| IQ: *M* (*SD*) | 109.66 (11.41) | 96.6 (12.63) | 93.4 (11.73) | 88.94 (13.21) |
| Reading: *M* (*SD*) | 108.7 (12.83) | 91.61 (17.43) | 86.51 (15.24) | 79.43 (15.37) |
| Maths: *M* (*SD*) | 115.52 (18.11) | 89.67 (18.04) | 82.73 (14.19) | 79.58 (14.16) |

A similar pattern was observed in the MRI sample, but there were fewer significant differences (see Tables 3, S8 & S9). The control group had significantly fewer diagnoses overall relative to each data-driven group. C1 had a significantly higher proportion of ADHD diagnoses relative to the comparison sample and C3. The comparison sample included a greater number of girls compared to C1, but the proportion of girls and boys did not significantly differ between the data-driven groups. As in the full sample, the comparison sample scored significantly higher than all of the data-driven groups on IQ, maths, and reading; and C1 scored significantly higher than C2 and C3 on IQ, maths, and reading. Mean functional connectivity did not differ between the groups but the comparison sample moved significantly less in the scanner compared to C1 and C2.

**Table 3.** Group Characteristics in the MRI sample.

|  | Controls | C1 | C2 | C3 |
| --- | --- | --- | --- | --- |
| *N* | 62 | 70 | 56 | 47 |
| Boys: *n* (%) | 28 (45.16%) | 55 (78.57%) | 33 (58.93%) | 26 (55.32%) |
| Girls: *n* (%) | 34 (54.84%) | 15 (21.43%) | 23 (41.07%) | 21 (44.68%) |
| Diagnosis: *n* (%) | 3 (4.84%) | 29 (41.43%) | 17 (30.36%) | 19 (40.43%) |
| ADHD: *n* (%) | 1 (1.61%) | 22 (31.43%) | 8 (14.29%) | 4 (8.51%) |
| Dyslexia: *n* (%) | 2 (3.23%) | 3 (4.29%) | 8 (14.29%) | 6 (8.51%) |
| Autism: *n* (%) | 0 (0%) | 8 (11.43%) | 1 (1.79%) | 4 (8.51%) |
| Age: *M* (*SD*) | 10.77 (2.05) | 10.71 (2.49) | 10.06 (1.77) | 10.26 (2.34) |
| IQ: *M* (*SD*) | 110.28 (10.57) | 100.32 (12.73) | 94.38 (11.17) | 89.35 (17.37) |
| Reading: *M* (*SD*) | 108.38 (11.69) | 95.06 (17.78) | 86.25 (15.44) | 78.35 (15.95) |
| Maths: *M* (*SD*) | 116.34 (20.32) | 94.87 (20.89) | 83.14 (13.31) | 79.96 (14.87) |
| Mean FC: *M* (*SD*) | 0.081 (0.027) | 0.083 (0.03) | 0.093 (0.028) | 0.08 (0.031) |
| Motion: *M* (*SD*) | 0.167 (0.076) | 0.226 (0.1) | 0.209 (0.092) | 0.207 (0.089) |

### 3.3. Global Connectome Properties

We examined the difference between the groups on global organisational properties of the functional connectome using ANCOVAs including age, gender, framewise displacement, and mean functional connectivity as nuisance covariates. There were no significant differences on any of the graph metrics analysed on the 25% group-thresholded connectome (see Table 4). Furthermore, there was no significant effect of group on any of the graph metrics when comparing the area under the curve (AUC) across all group-thresholds to a distribution of 1000 permutations where group labels were randomly shuffled (all *p* > 0.102; see Table S10). Similar results were also obtained for the individually thresholded connectomes, which may be more sensitive to individual differences in global organisation. There were no significant group differences in any of the graph metrics at the 25% threshold (see Table 5) and or across all thresholds when examining the AUC (all *p* > ###; see Table S11). In sum, there was no evidence that the groups significantly differed in global brain organisation.

**Table 4.**
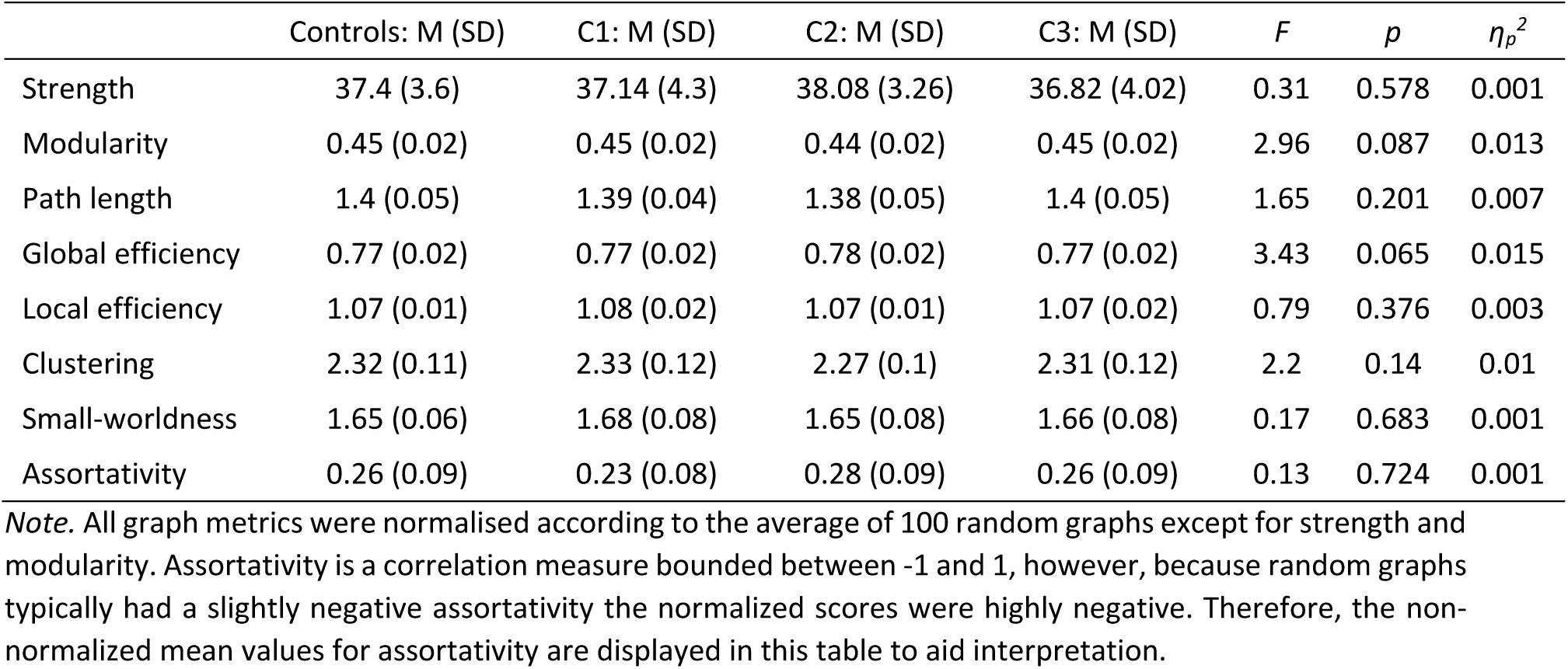
Group differences in global graph metrics at the 25% group threshold.

**Table 5.** Group differences in global graph metrics at the 25% individual threshold.

| | Controls: M (SD) | C1: M (SD) | C2: M (SD) | C3: M (SD) | *F* | *p* | $\eta_p^2$ |
| --- | --- | --- | --- | --- | --- | --- | --- |
| Strength | 49.93 (4.49) | 50.66 (4.76) | 51.75 (4.8) | 49.86 (4.05) | 0.08 | 0.774 | <0.001 |
| Modularity | 0.36 (0.04) | 0.36 (0.03) | 0.34 (0.04) | 0.36 (0.03) | 1.78 | 0.184 | 0.008 |
| Path length | 1.24 (0.12) | 1.23 (0.1) | 1.22 (0.1) | 1.23 (0.07) | 0.11 | 0.74 | <0.001 |
| Global efficiency | 0.85 (0.08) | 0.86 (0.07) | 0.86 (0.07) | 0.85 (0.05) | 0.04 | 0.845 | <0.001 |
| Local efficiency | 1.06 (0.15) | 1.06 (0.12) | 1.06 (0.13) | 1.05 (0.09) | 0.46 | 0.5 | 0.002 |
| Clustering | 1.87 (0.41) | 1.85 (0.37) | 1.82 (0.41) | 1.82 (0.26) | 0.91 | 0.343 | 0.004 |
| Small-worldness | 1.54 (0.44) | 1.53 (0.38) | 1.51 (0.44) | 1.49 (0.27) | 0.89 | 0.346 | 0.004 |
| Assortativity | 0.29 (0.09) | 0.28 (0.08) | 0.3 (0.09) | 0.29 (0.07) | 0.13 | 0.724 | 0.001 |

*Note.* All graph metrics were normalised according to the average of 100 random graphs except for strength and modularity. Assortativity is a correlation measure bounded between -1 and 1, however, because random graphs typically had a slightly negative assortativity the normalized scores were highly negative. Therefore, the non-normalized mean values for assortativity are displayed in this table to aid interpretation.

### 3.4. Intrinsic Connectivity Networks

We examined the relationship between ICN connectivity and the behavioural profiles using Partial Least Squares (PLS) regression. Cross-validated prediction error was assessed using a casewise calculation of the root mean square error, which is directly proportional to classification accuracy and also provided for interpretability. The lowest prediction error was achieved with three PLS components (RMSE = 0.466, SEM = 0.019; Accuracy = 0.341, SEM = 0.027). To find a balance between accuracy and parsimony we selected the simplest model within one standard error of the lowest cross-validated RMSE (Hastie et al., 2009). In this case, the prediction error in models with fewer components was greater than one standard error above the RMSE with three components. We then compared this model’s accuracy with three components to that would be expected by chance when group labels were randomly shuffled 1000 times. The three components of ICN connectivity significantly explained group membership relative to the permuted null models (permuted RMSE = 0.519, SEM = 0.019, *p* = 0.005). This finding was reproduced over different connectome thresholds (see Table S12) and was significant across all thresholds when computing the AUC (AUC = 2.363, permuted AUC = 2.594, SEM = 0.087, *p* = 0.007).

We then assessed the contribution of specific ICN connections at the 25% threshold by examining the loadings of a PLS model with three components fit to 1000 bootstrapped samples. Loadings were aligned across the bootstrapped samples using an orthogonal Procrustes rotation and component scores were re-computed. We then tested whether group differences in component scores significantly differed from chance when group labels were permuted 1000 times. The top 25% of loadings and group differences are shown in in Figure 2). The highest loadings for PLS1 were ICN connections with the dorsal attention and limbic networks. Group C2 scored significantly higher on this component compared to C1 (*p* < 0.001) and Controls scored marginally higher than C1 but this was borderline significant (*p* = 0.058). The highest loadings for PLS2 were widespread across all eight ICNs. Controls generally scored higher on this component compared to the other groups but this was only statistically significant for the comparison with C2 (*p* = 0.012, other *p’*s > 0.073). Finally, highest loadings for PLS3 included connections with the subcortical, default-mode, and dorsal attention networks. C3 generally scored lower on this component compared to the other groups but these differences were not statistically significant (*p* > 0.093).

**Figure 2.**
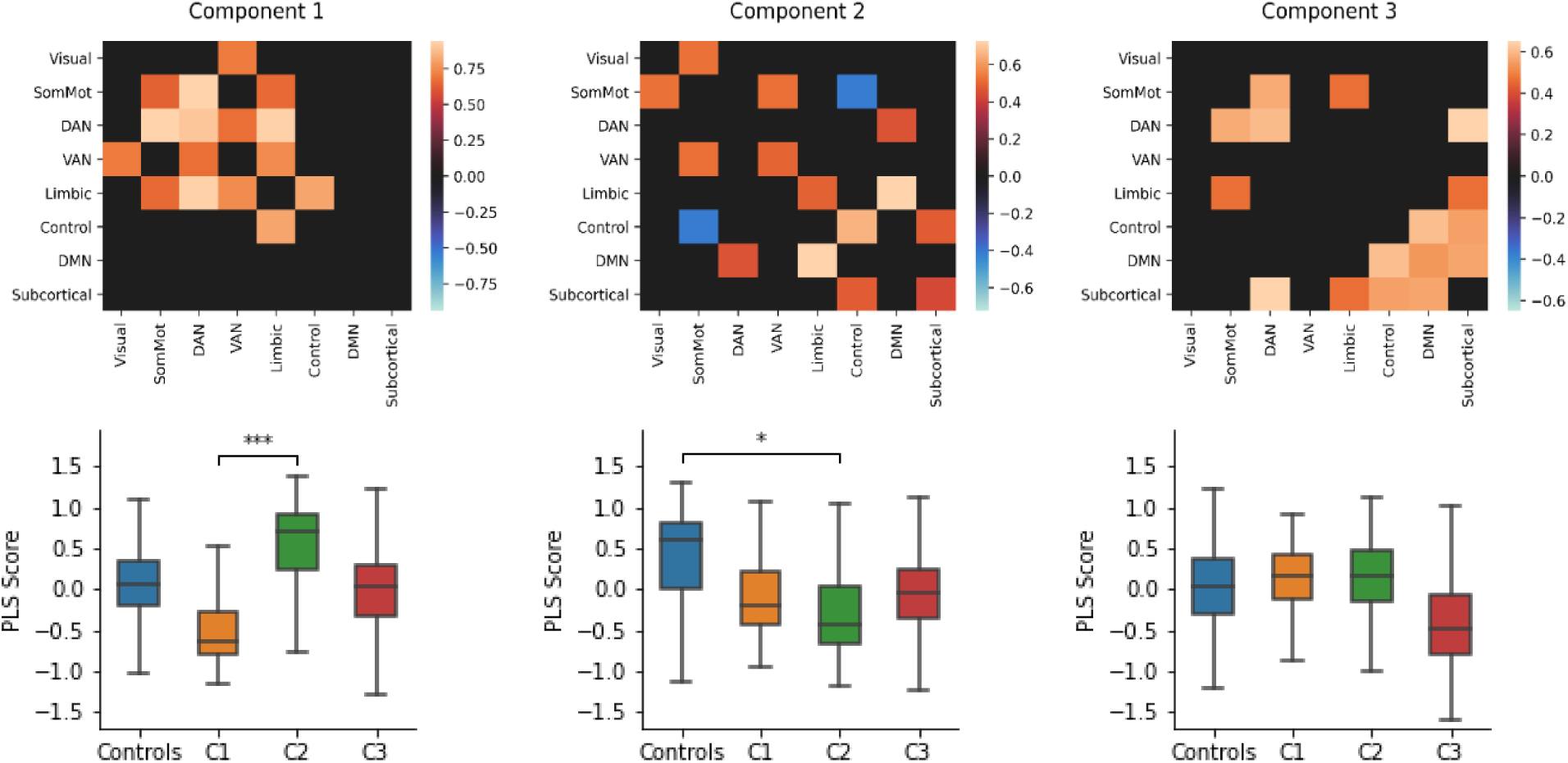
PLS components of ICN connectivity that predicted group membership. The heatmaps (top) show the 25% largest loadings of ICN connections onto each component relative to their standard error over 1000 bootstrapped samples. The boxplots (bottom) show the bootstrapped distribution of component scores for each group and significant group differences assessed by a permutation test. ICN abbreviations: Somatomotor (SomMot), dorsal attention network (DAN), ventral attention network (VAN), fronto-parietal (Control), and default mode network (DMN). ***p<0.001, *p<0.05

### 3.5. Nodal Strength

PLS applied to nodal strength across the entire functional connectome also distinguished held-out children’s behavioural profile better than chance. The lowest prediction error was achieved with 10 PLS components (RMSE = 0.466, SEM = 0.02; Accuracy = 0.341, SEM = 0.029); however, a simpler model with 5 components was within one standard error (RMSE = 0.481, SEM = 0.016; Accuracy = 0.319, SEM = 0.022). This simpler model significantly explained group membership compared to a null model based on 1000 shuffled samples (permuted RMSE = 0.519, SEM = 0.018, *p* = 0.021). This finding was reproduced over multiple connectome thresholds (see Table S13) and significant across all thresholds when computing the AUC (AUC = 2.387, permuted AUC = 2.591, SEM = 0.074, *p* = 0.003).

The contribution of specific nodes to the PLS components and group differences were analysed as for ICNs. Group differences were only found for the first two components, which are displayed in Figure 3 (see Figure S7 for components 3-5). The top 5% of loadings for PLS1 predominantly included medial regions of the default-mode, limbic, visual, and fronto-parietal network, as well as the bilateral hippocampi. Controls scored significantly higher than C2 (*p* = 0.004), and C1 scored higher than C2 (*p* < 0.001) and C3 (*p* = 0.004). The highest loadings from PLS2 predominantly included lateral regions of the default-mode network, as well as lateral temporal regions of the limbic and fronto-parietal networks. Controls scored marginally higher on PLS2 compared to C3, but this was borderline significant (*p* = 0.056).

**Figure 3.**
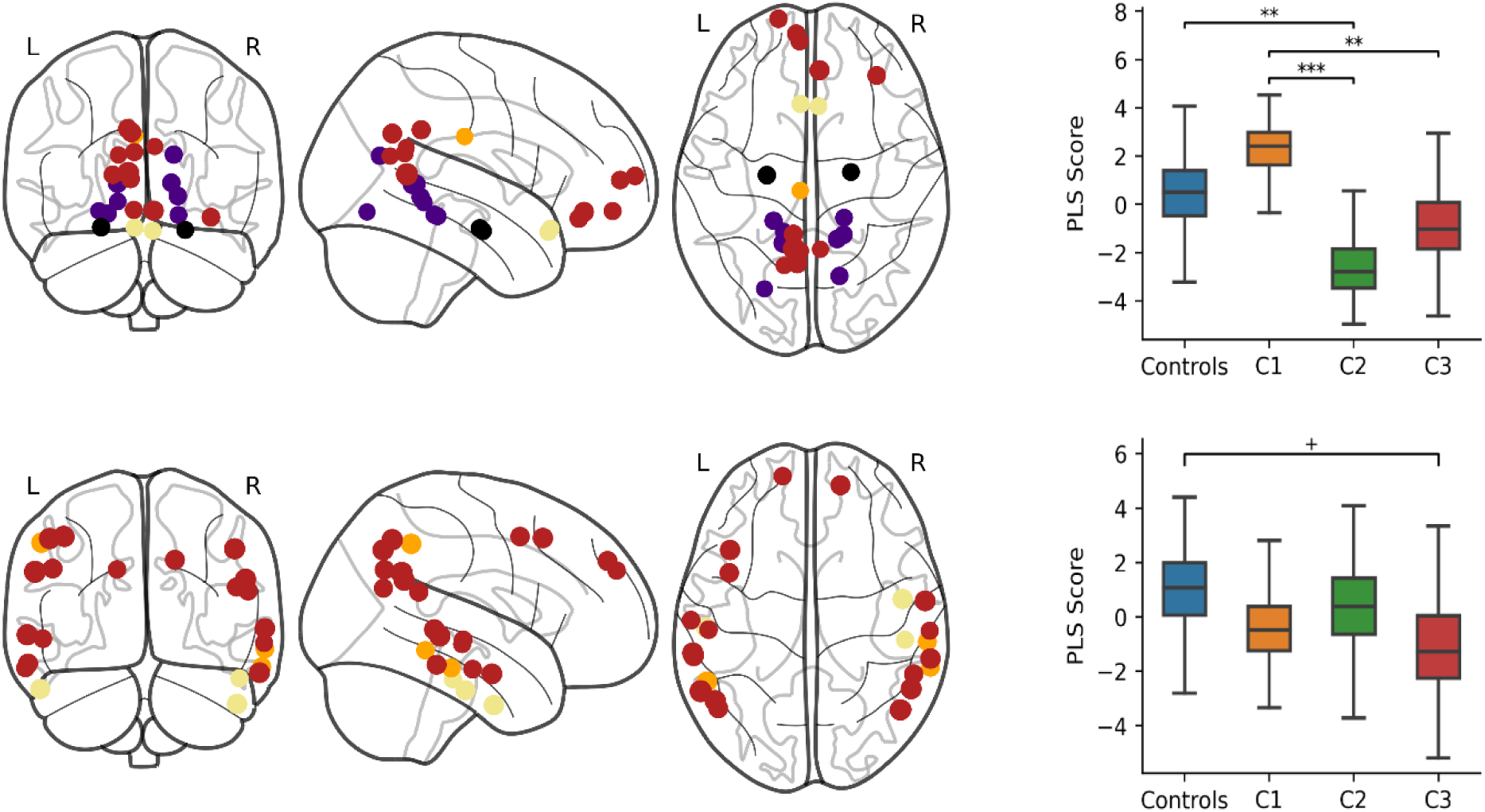
PLS components of nodal strength that predicted group membership. The brain plots (left) show the 5% largest loadings of nodes onto each component (PLS1 top, PLS2 bottom) relative to their standard error over 1000 bootstrapped samples. The size of the node is proportional to its absolute loading and the colour corresponds to its ICN: default-mode (red), limbic (cream), visual (purple), fronto-parietal (orange), and subcortical (black). The boxplots (right) show the bootstrapped distribution of component scores for each group and significant group differences assessed by a permutation test. ***p<0.001, **p<0.01, ^+^p<0.06

**Figure 4.**
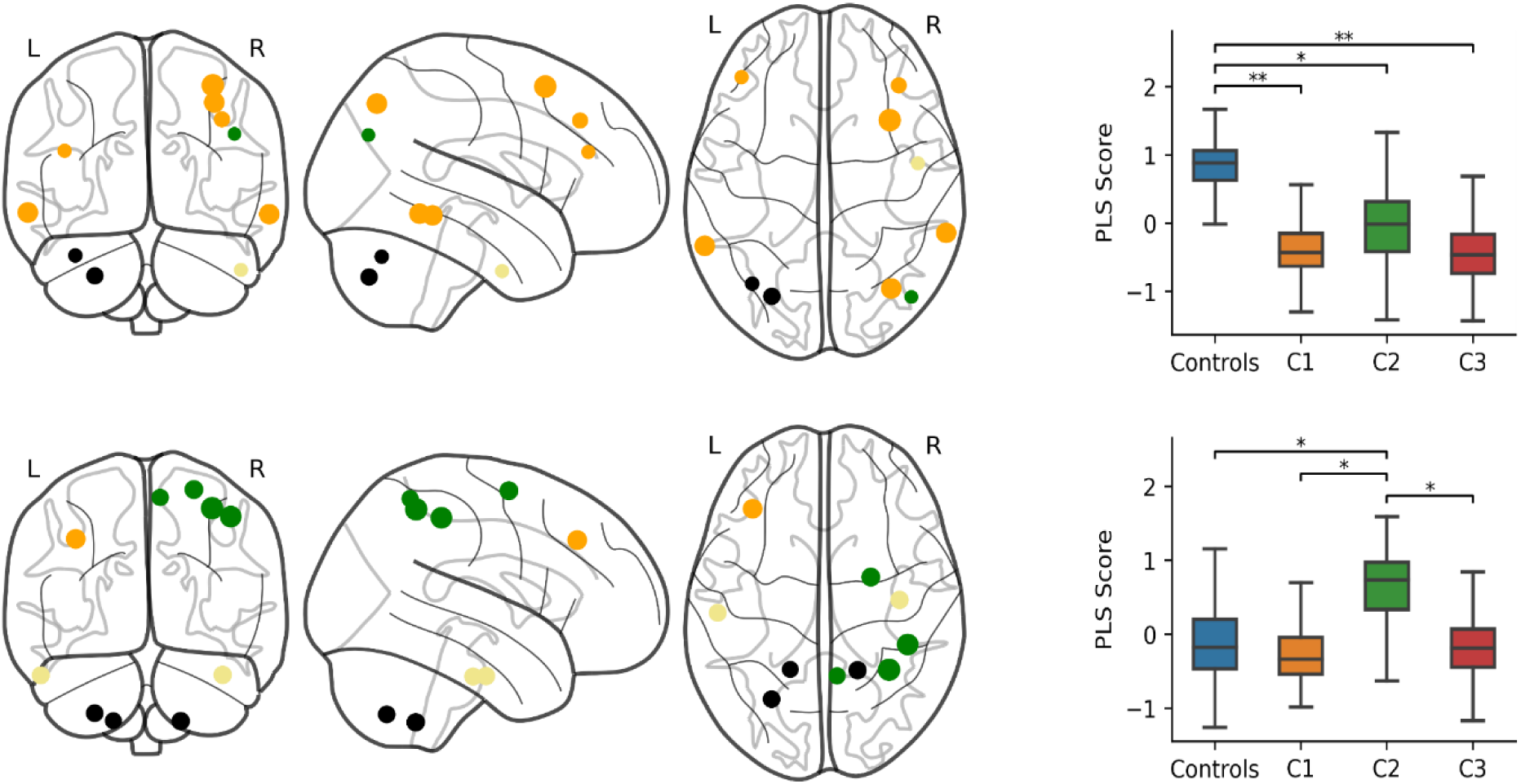
PLS components of connector hub strength that predicted group membership. The brain plots (left) show the 30% largest loadings of connector hubs onto each component (PLS1 top, PLS2 bottom) relative to their standard error over 1000 bootstrapped samples. The size of the node is proportional to its absolute loading and the colour corresponds to its ICN: fronto-parietal (orange), subcortical (black), dorsal attention (green), and limbic (cream). The boxplots (right) show the bootstrapped distribution of component scores for each group and significant group differences assessed by a permutation test. **p<0.01, *p<0.05

### 3.6. Hubs

We identified 31 connector hubs, defined as the top 30^th^ percentile for both betweenness centrality and participation coefficient. Connector hubs included regions within the dorsal attention, fronto-parietal, cerebellar and limbic networks. The PLS analysis demonstrated that the strength of connector hubs significantly predicted group membership for held-out data. The most accurate and parsimonious model included two PLS components (RMSE = 0.472, SEM = 0.015; Accuracy = 0.322, SEM = 0.021) and significantly explained group membership above a null model based on 1000 shuffled samples (permuted RMSE = 0.516, SEM = 0.018, *p* = 0.011). This finding was reproduced over multiple connectome thresholds (see Table S14) and significant across all thresholds when computing the AUC (AUC = 2.353, permuted AUC = 2.564, SEM = 0.077, *p* = 0.002).

The top 30% of loadings onto PLS components and group differences are displayed in Figure PLS1 largely included bilateral regions of the fronto-parietal network as well as the left cerebellum, right temporal pole of the limbic network, and right intraparietal sulcus of the dorsal attention network. Controls scored significantly higher on PLS1 compared to C1 (*p* = 0.004), C2 (*p* = 0.034), and C3 (*p* = 0.002). The highest loadings for PLS2 included the right dorsal attention network, bilateral cerebellum, bilateral temporal poles of the limbic network, and the left lateral prefrontal cortex of the fronto-parietal network. C2 scored significantly higher on PLS2 compared to Controls (*p* = 0.034), C1 (*p* = 0.006), and C3 (*p* = 0.044).

We also identified 103 provincial hubs in the top 30^th^ percentile for within module strength that had a participation coefficient in the bottom 70^th^ percentile. Provincial hubs largely included regions in the default mode, somatomotor, visual, ventral attention and frontoparietal networks, as well as the thalamus. The PLS analysis demonstrated that the strength of provincial hubs significantly predicted group membership for held-out data. The most accurate and parsimonious model included seven PLS components (RMSE = 0.469, SEM = 0.017; Accuracy = 0.336, SEM = 0.023) and significantly explained group membership above a null model based on 1000 shuffled samples (permuted RMSE = 0.522, SEM = 0.019, *p* = 0.003). This finding was reproduced over different connectome thresholds (see Table S15) and significant across all thresholds when computing the AUC (AUC = 2.368, permuted AUC = 2.6, SEM = 0.081, *p* = 0.005).

Significant group differences were only observed on the first component, which is displayed in Figure 5 with component two (see Figure S8 for components 3-7). The top 10% of loadings for PLS1 largely included regions of the default-mode network, as well as the bilateral lingual gyri of the visual network and right orbitofrontal cortex of the limbic network. C1 scored significantly higher on PLS1 compared to C2 (*p* = 0.012) and C3 (*p* = 0.008), and Controls scored significantly higher than C3 (*p* = 0.030) and marginally higher than C2, however this was borderline significant (*p* = 0.054). The highest loadings for PLS2 included bilateral lateral temporal and parietal regions of the default-mode network, bilateral regions of the fronto-parietal network, the parietal operculum of the somatomotor network, and the left temporal pole of the limbic network. Controls scored higher on PLS2 than C1 (*p* = 0.102) and C3 (*p* = 0.098) but these differences were not statistically significant.

**Figure 5.**
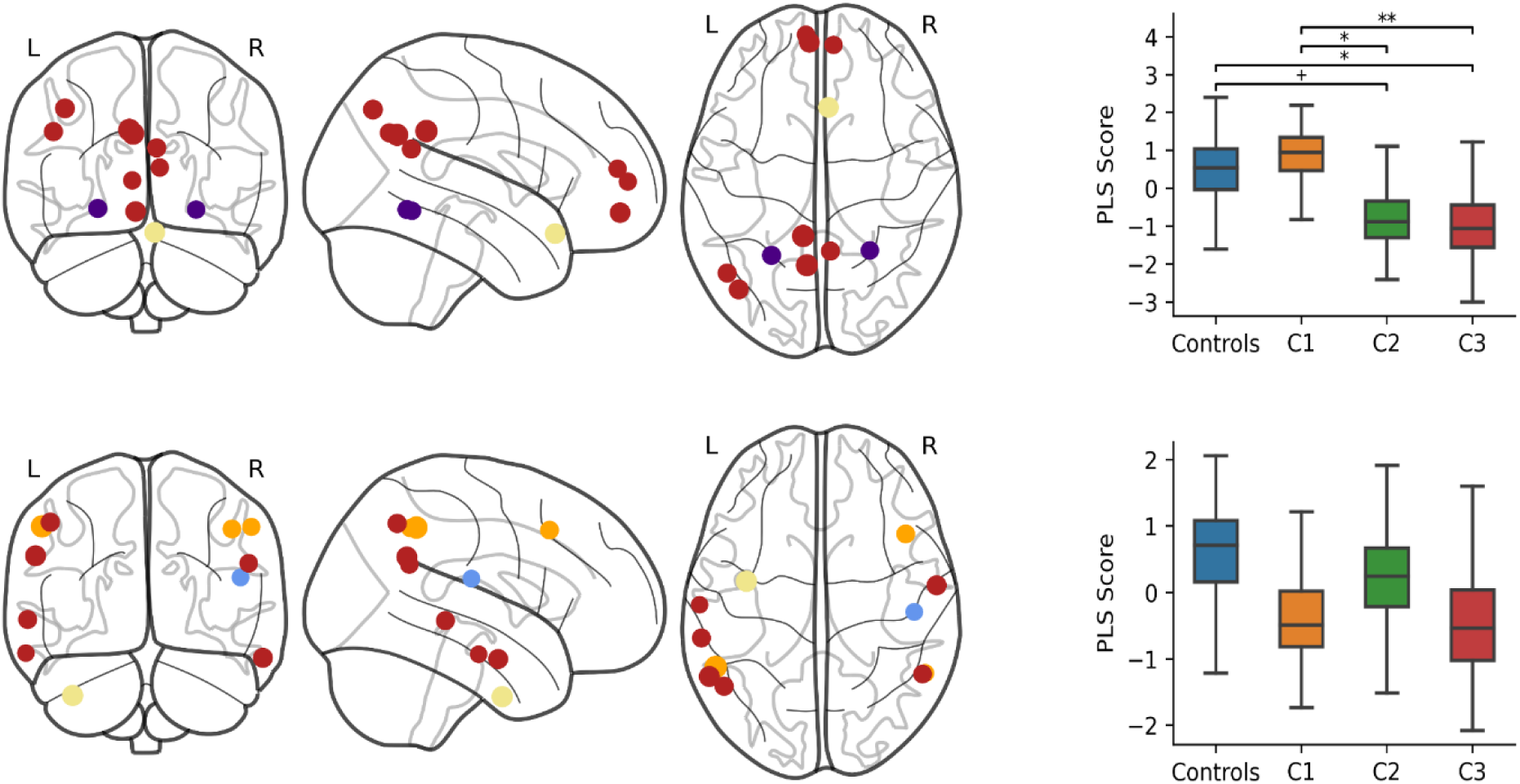
PLS components of provincial hub strength that predicted group membership. The brain plots (left) show the 10% largest loadings of provincial hubs onto each component (PLS1 top, PLS2 bottom) relative to their standard error over 1000 bootstrapped samples. The size of the node is proportional to its absolute loading and the colour corresponds to its ICN: default-mode (red), visual (purple), limbic (cream), fronto-parietal (orange), and somatomotor (blue). The boxplots (right) show the bootstrapped distribution of component scores for each group and significant group differences assessed by a permutation test. **p<0.01, *p<0.05, ^+^p<0.06

## 4. Discussion

We tested whether a large mixed sample of children at neurodevelopmental risk could be grouped according to their behavioural profiles. Consensus community detection within a network analysis identified three groups with distinct behavioural profiles. These groups were characterised by principal difficulties with hot executive function (C1), cool executive function (C2), and learning (C3). Next, we created functional connectomes for a subset of these children and tested whether group membership could be predicted by different aspects of functional brain organisation. Whilst there were no group differences in global organisational properties, multivariate patterns of connectivity at the level of ICNs, nodes, and hubs significantly predicted group membership in held-out data.

### 4.1. Behavioural profiles

Ratings of inattention, cool executive function difficulties, and learning problems were in the ‘elevated’ or ‘very elevated’ range for all groups and significantly higher than the comparison sample. However, the degree of difficulties in specific behavioural domains were distinct between the three behavioural profiles capturing principal difficulties with hot executive function (C1), cool executive function (C2), and learning problems (C3). These profiles were largely consistent with previously identified profiles in a smaller sample from the same cohort of children (Bathelt, Holmes, et al., 2018).

The largest group, C1, were uniquely characterised by ratings of aggressive behaviour in the very elevated range, which exceeded all other groups. This was accompanied by the highest ratings of hyperactivity/impulsivity and poor peer relationships. Within the wider literature these difficulties seem to co-occur and are more related to tasks tapping hot executive function, particularly emotion regulation and impulse control (Zelazo & Carlson, 2012). High rates of peer relation difficulties are not surprising in this group given the robust association between emotion regulation and prosocial behaviour in children (Eisenberg et al., 2007; Hastings et al., 2005; Liew et al., 2011; Moriguchi et al., 2020), and links between behavioural difficulties and pragmatic language (Hawkins et al., 2016; Ketelaars et al., 2010), which together may influence how these children form social networks. Although hot and cool executive function are often closely associated (Zelazo & Carlson, 2012), this group had the lowest ratings of inattention and cool executive function difficulties, and their IQ was age-appropriate. C1 also had the lowest ratings of learning problems and they performed significantly higher on the maths and reading tasks compared to the other data-driven profiles. Despite relatively fewer cognitive difficulties, this group included the highest proportion of overall diagnoses, ADHD, and autism relative to some of the data-driven profiles, and significantly less children with dyslexia than expected compared to C2 and C3. Approximately one third of children in C1 had a diagnosis of ADHD, but the relatively high occurrence of autism in this group, age-appropriate IQ (Frazier et al., 2004), and numerous ADHD diagnoses in the other two groups suggests that this group does not simply reflect ADHD. This group appear to be primarily characterised by behavioural difficulties, rather than cognitive difficulties. C1 also included a greater proportion of boys than all other groups. Previously, boys have generally been shown to have greater externalising and social difficulties than girls in this sample (J. Holmes, Mareva, et al., 2020) and the higher rates of neurodevelopmental diagnoses in boys is consistent with the wider literature (e.g. Russell et al., 2014). Within the context of this particular cohort, C1 appears to capture those with the most pronounced difficulties in hot executive function.

In contrast, C2 had relatively low ratings of aggression and peer relation difficulties. C2 were instead characterised by particularly elevated behavioural difficulties with cool executive function. They were rated significantly higher on inattention and (cool) executive function difficulties compared to all other groups. This suggests that these children particularly struggle with the cognitive control needed to concentrate, redirect their attention, plan, and organise (Diamond, 2013). Although it should be noted that these children also scored in the very elevated range for hyperactivity/impulsivity. This profile is somewhat similar to the characteristic symptoms of combined type ADHD, yet less than a fifth of children were diagnosed with ADHD in the group and it was more common in C1. C2 had moderate difficulties on tasks tapping general cognitive ability and academic achievement, performing better than children in C3 but worse than those in C1. Thus, C2 appears to capture those with the most pronounced difficulties in cool executive function behaviours, with moderate IQ and learning difficulties.

The smallest group, C3, were characterised by the highest ratings of learning problem behaviours, which significantly exceeded the other two groups. Their behavioural difficulties with cognitive control were comparable to C1; however, ratings of hyperactivity and aggression were within the normal range. In addition, children in C3 performed the most poorly on tasks tapping IQ and academic achievement. This group may represent a smaller proportion of the population with more selective learning and cognitive difficulties with relatively low rates of hyperactivity and conduct problems. Elevated difficulties with peer relationships in this group is perhaps more surprising; however, peer rejection is more common in children with learning difficulties (Frederickson & Furnham, 2004; Parhiala et al., 2015; Pijl & Frostad, 2010; Siperstein et al., 2007; Wiener & Schneider, 2002). There are many potential causes of peer problems in children with learning difficulties including cognitive difficulties, stigma, social anxiety, and victimization for being enrolled in additional education programs (Livingston et al., 2018). In sum, C3 appears to capture those with the most pronounced cognitive and learning difficulties.

### 4.2. Functional brain organisation

Despite large differences in children’s behavioural profiles, no group differences in functional brain organisation were identified on a global level. These findings were consistent for both individual and group thresholds across a range of values. This suggests that across our diverse sample behavioural difficulties are not well explained by global differences in functional organisation across the whole brain, at least in so far as we were able to capture them here. Network organisation is similarly modular, clustered, efficient, and assortative across the groups. Instead, behavioural profiles were significantly predicted by specific multivariate patterns of functional connectivity between ICNs, nodal strength, and hub strength. These associations were reproduced across multiple connectome thresholds and significant across all thresholds when considering the area under the curve.

#### 4.2.1. Comparison sample vs data-driven groups

Some of the most apparent differences in functional organisation characterise all three of our data-driven groups, relative to the comparison sample. Comparison children scored significantly higher on the first PLS component of connector hub strength, compared to all other groups. This first PLS component predominantly loaded on to bilateral regions of the fronto-parietal network, which has a critical role in integrating information between networks to initiate and regulate cognitive control (Astle, Luckhoo, et al., 2015; Cole et al., 2015, 2017; Marek & Dosenbach, 2018; Sheffield et al., 2015). Connector hubs play an important role in global integration: as networks specialise and segregate in childhood development, hubs become increasingly structurally connected both within and between networks, maintaining efficient communication across the connectome and supporting the typical development of cool executive function (Baum et al., 2017). This distinction in fronto-parietal connector hubs between comparison children and all other groups suggests that this is a relatively generic feature of children at neurodevelopmental risk, not related to specific profiles of behavioural difficulties. That said, behavioural difficulties associated with cognitive control and learning were consistently elevated in all groups relative to the comparison sample. This finding mirrors a result from a recent study of the same cohort, which demonstrated that children with no or selective cognitive difficulties have more highly connected structural hubs (Siugzdaite et al., 2020). Structural connector hubs have also been shown to have a particularly important role in predicting academic progress in children from the same cohort (Bathelt, Gathercole, et al., 2018). Taken together, this suggests that connector hubs play a key and relatively non-specific role in distinguishing children at neurodevelopmental risk from comparison samples. This effect is remarkably consistent across the structural and functional connectome despite the differences in modality, pre-processing, parcellation, analysed sample, and analysis method. In sum, this suggests that emerging neurodevelopmental differences in connector hub structural connectivity may have consequences for hub function and cognitive development.

#### 4.2.2. Distinguishing children with hot and cool executive function difficulties

The PLS of ICN connectivity and nodal strength primarily distinguished C1 and C2 – those with relatively greater difficulties in hot versus cool executive function, respectively. In fact, the first component of ICN connectivity only significantly differed between these two behavioural profiles. Notably, connections of the dorsal attention and limbic networks loaded most strongly on this component. The dorsal attention network is commonly implicated in tasks requiring cognitive control, such as top-down attention and working memory (Rottschy et al., 2012; Vossel et al., 2014), and training working memory in childhood has been shown to increase functional connectivity within this network (Astle, Barnes, et al., 2015). Connectivity of the limbic network, including regions such as the orbitofrontal cortex, is particularly associated with hot executive function (Ho et al., 2015; Hulvershorn et al., 2014; Karalunas et al., 2014; Posner et al., 2014). Children in C2 had positive scores on this component suggesting that over-connectivity between these networks may be associated with particularly elevated cognitive difficulties, whereas children in C1 had negative scores suggesting that under-connectivity may indicate particularly elevated difficulties with emotional control.

A similar distinction was observed for the first component of nodal strength, where children in C1 scored significantly higher than those in C2 and C3, though this distinction was greater for children in C2 who had highly negative scores. This component included high loadings from medial default-mode regions, the bilateral orbitofrontal cortex, bilateral hippocampi, and visual association areas. Strikingly, the same group distinction in a smaller sample of this cohort revealed a highly similar pattern of structural connectivity differences, which were localised to the orbitofrontal cortex, anterior cingulate, medial temporal lobe, visual cortex, and basal ganglia (Bathelt, Holmes, et al., 2018). The orbitofrontal cortex is important in hot executive function, emotion behaviour, and value-based decision-making (Fuster, 2001; Gazzaniga et al., 2014; Padoa-Schioppa & Conen, 2017). It is strongly connected to the hippocampus and amygdala (Cavada et al., 2000; Chudasama & Robbins, 2006; Morecraft et al., 1992; Zald et al., 2014), and functional connectivity between these regions is associated with poorer emotional control (Ho et al., 2015), mood problems (Hulvershorn et al., 2014; Posner et al., 2014), and temperament difficulties in childhood (Karalunas et al., 2014). Furthermore, data-driven subtyping of functional connectivity in this cortical-subcortical network has independently provided evidence for hot and cool executive function subgroups of children (Costa Dias et al., 2015). In the current sample, structural and functional connectivity of the orbitofrontal cortex, anterior cingulate, and medial prefrontal cortex were implicated in this group distinction. These regions contribute to the anterior default-mode network, which is implicated in emotion processing, self-referential thought, and social cognition (Raichle, 2015; Schilbach et al., 2008). These links to emotion regulation and social skills are notable considering the pronounced difficulties that children in C1 experienced in these domains.

The medial default-mode network was further implicated in a distinction between the hot (C1) and cool executive function subgroups (C2, C3) on the first component of provincial hub strength. Specifically, C2 and C3 showed under-connectivity in these provincial hubs relative to C1 and the comparison sample. These regions are particularly important for integrating information within the default-mode network and previous work in the same cohort has shown that functional connectivity within the default-mode network is related to underlying structural connectivity differences in the cingulum (Bathelt et al., 2019). Interestingly, in this previous work the relationship between structure and function was only apparent for children with poor cognitive ability, such as those in C2 and C3, suggesting that variability in cingulum structural connectivity may only have significant functional consequences at the lower end of the spectrum. Alterations in default mode network connectivity have also been widely documented in a range of neurodevelopmental and mental health conditions (Menon, 2011). For example, altered connectivity between the default mode network and externally-oriented task-positive networks has been linked to executive function difficulties (Abbott et al., 2016), inattention and hyperactivity/impulsivity (Cai et al., 2018; H. Lin et al., 2018; Sripada et al., 2014). Our results do not distinguish which networks these default-mode regions are under-connected to, but they instead demonstrate that these regions are generally less well connected. This transdiagnositic feature of functional connectivity in neurodevelopmentally at-risk children may extend beyond specific inter-network connections evidenced in prior research and highlight an altered functional role of these regions in the whole connectome.

#### 4.2.3. Specific Group Distinctions

Evidence for individual group distinctions was also observed. C2 was significantly distinguished from all other groups on the second component of connector hub strength, which particularly included high loadings on the right dorsal attention network. C2 demonstrated over-connectivity in these regions and primarily differed from other groups in their degree of difficulties with inattention and cool executive function. It is possible that poor cognitive control in this group may be related to over-connectivity of dorsal attention connector hubs, for example to the default-mode network (H. Lin et al., 2018). There was more limited evidence that C3 were distinct from all other groups, which could suggest that the neurobiological correlates of this group with pronounced and more selective learning difficulties were more heterogeneous. However, C3 was partially distinguished from the other groups on the third component of ICN connectivity, which included high loadings on connections of the subcortical, default-mode, and dorsal attention networks. Group differences on this component were not statistically significant, but this extra component increased prediction accuracy of behavioural profiles on held-out data.

### 4.3. Limitations

There are several limitations to the current work. First, we note that the comparison sample moved less during scanning than the other groups. We took many steps to control motion artefacts at the individual and group level. This included a thorough assessment of different resting-state fMRI pre-processing techniques, exclusion of high motion participants, censoring of high motion volumes, physiological and motion confound regression, and the inclusion of motion and mean functional connectivity in group-level analyses. Second, we excluded approximately a third of resting-state scans due to high movement. While this was necessary to ensure data quality, it may limit statistical power and may have excluded children who were younger, particularly anxious, or hyperactive. Importantly, however, the behavioural profiles were very similar in both the full sample and MRI sample. Third, while the Conners questionnaire is well-validated and used clinically, it does not cover all relevant aspects of children’s cognitive, emotional, and social behaviour. Additionally, this could lead to some bias in the composition of the data-driven groups. Fourth, we assessed correlations in the behavioural network; this measures how similar each child’s behavioural profile is to one another but disregards overall differences in severity of difficulties. Distance metrics that satisfy triangular inequality would need to be used to take into account differences in overall severity. Fifth, whilst held-out children’s behavioural profile could be predicted above chance level on the basis of their functional connectivity, these predictions were not sufficiently accurate to warrant use of these methods in an applied setting.

### 4.4. Conclusion

We identified distinct data-driven behavioural profiles that transcend diagnostic categories in a large heterogeneous sample of children at neurodevelopmental risk. These groups were not associated with differences in global organisation of brain function, but were associated with multivariate patterns of connectivity between ICNs, nodes, and hub regions. Children with more pronounced hot or cool executive function difficulties were distinguished by connectivity in ICNs implicated in cognitive control, emotion processing, and social cognition. Furthermore, all of the data-driven groups differed from the comparison sample in connectivity of the fronto-parietal connector hubs. Our findings suggest both specific and more general neurodevelopmental risk factors in the functional connectome, which corroborate with previously reported risk factors in the structural connectome.

## Data Availability

Access to this dataset is currently managed by the Centre for Attention Learning and Memory (CALM) Management Committee. It is currently being prepared for external access. However, the analysis code can be found in the OSF repository.

https://osf.io/cvsu2/

## 5. Acknowledgments

The authors were supported by the Medical Research Council program grant MC-A0606-5PQ41. We would like to thank all members of the CALM Team for their help with recruitment, data collection, and data management, as well as all of the children and parents for their participation in the study. The CALM Team includes lead investigators Duncan Astle, Kate Baker, Susan Gathercole, Joni Holmes, Rogier Kievit and Tom Manly. Data collection is assisted by a team of researchers and PhD students that includes Danyal Akarca, Joe Bathelt, Marc Bennett, Giacomo Bignardi, Sarah Bishop, Erica Bottacin, Lara Bridge, Diandra Brkic, Annie Bryant, Sally Butterfield, Elizabeth Byrne, Gemma Crickmore, Edwin Dalmaijer, Fánchea Daly, Tina Emery, Laura Forde, Grace Franckel, Delia Furhmann, Andrew Gadie, Sara Gharooni, Jacalyn Guy, Erin Hawkins, Agnieszka Jaroslawska, Sara Joeghan, Amy Johnson, Jonathan Jones, Silvana Mareva, Elise Ng-Cordell, Sinead O’Brien, Cliodhna O’Leary, Joseph Rennie, Ivan Simpson-Kent, Roma Siugzdaite, Tess Smith, Stephani Uh, Maria Vedechkina, Francesca Woolgar, Natalia Zdorovtsova, Mengya Zhang. The authors wish to thank the many professionals working in children’s services in the South-East and East of England for their support, and to the children and their families for giving up their time to visit the clinic. We would also like to thank the radiographers who support the excellent paediatric scanning at the MRC CBSU

## Notes

### Competing Interest Statement

The authors have declared no competing interest.

### Author Declarations

Ethical approval for the Centre for Attention Learning and Memmory cohort study was granted by the National Health Service (NHS) Health Research Authority NRES Committee East of England, REC approval reference 13/EE/0157, IRAS 127675.

