## Supplementary materials for "A Transdiagnostic Data-driven Study of Children’s Behaviour and the Functional Connectome"

### Developmental Cognitive Neuroscience

### Age Distributions

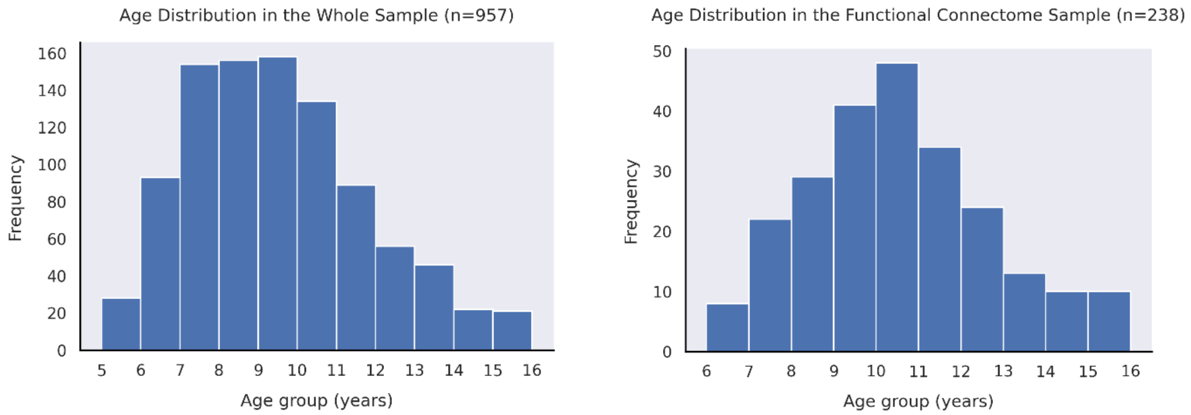


**Figure S1**. Age distributions in the whole sample (left) and the functional connectome sample (right).

### MRI Sample Demographics

#### Table S1. MRI Sample Characteristics

| | Struggling learners (*n*=175) | Comparison sample (*n*=63) |
|---|---|---|
| Age in years: *M* (*SD*) | 10.36 (2.23) | 10.79 (2.04) |
| Boys: *n* | 115 (65.71%) | 29 (46.03%) |
| Girls: *n* | 60 (34.29%) | 34 (53.97%) |
| No diagnosis: *n* | 109 (62.29%) | 60 (95.24%) |
| ADHD: *n* | 35 (20%) | 1 (1.59%) |
| Suspected ADHD: *n* | 10 (5.71%) | 0 (0%) |
| Autism: *n* | 13 (7.43%) | 0 (0%) |
| Dyslexia: *n* | 17 (9.71%) | 2 (3.17%) |

### Evaluation of rsfMRI denoising strategies

A number of pipelines were evaluated to denoise motion and physiological artefacts from the resting-state fMRI data using the fmridenoise package in Python (https://github.com/compneuro-ncu/fmridenoise). The pipelines included combinations of different methods and regressors:

- 24 Head Motion Parameters (24HMP) – Regression of the 6 rigid body realignment parameters, their squares, their first derivatives, and the squares of the first derivatives
- 8 Physiological regressors (8Phys) – Regression of the time series activity from the CSF and WM masks, their squares, their first derivatives, and the squares of the first derivatives
- 4 Global Signal Regressors (4GSR) – Regression of the whole-brain time series activity, its square, first derivative, and square of the first derivative
- Motion spike regression (SpikeReg) – Regression of volumes where framewise displacement (FD) was greater than 0.5mm or where the BOLD signal change (DVARS) was 3 standard deviations away from the mean (Power, Barnes, Snyder, Schlaggar, & Petersen, 2012)
- Anatomical component correction (aCompCor) – Regression of 10 principal components estimated from the CSF and WM matter masks (Behzadi, Restom, Liau, & Liu, 2007)
- ICA-AROMA – Estimation of independent components using MELODIC in FSL, automatic identification of noise components, and their subsequent regression from the data (Pruim et al., 2015)

24HMP_aCompCor_SpikeReg (coloured pink in the following figures) was selected because it was the best performing pipeline across a number of quality control measures. It is also widely implemented as the default denoising pipeline in Conn (Whitfield-Gabrieli & Nieto-Castanon, 2012). Although GSR pipelines performed well on various quality metrics, we did not implement it here because GSR is a highly controversial technique that may remove neural signal and introduce spurious anti-correlations (Murphy & Fox, 2017).

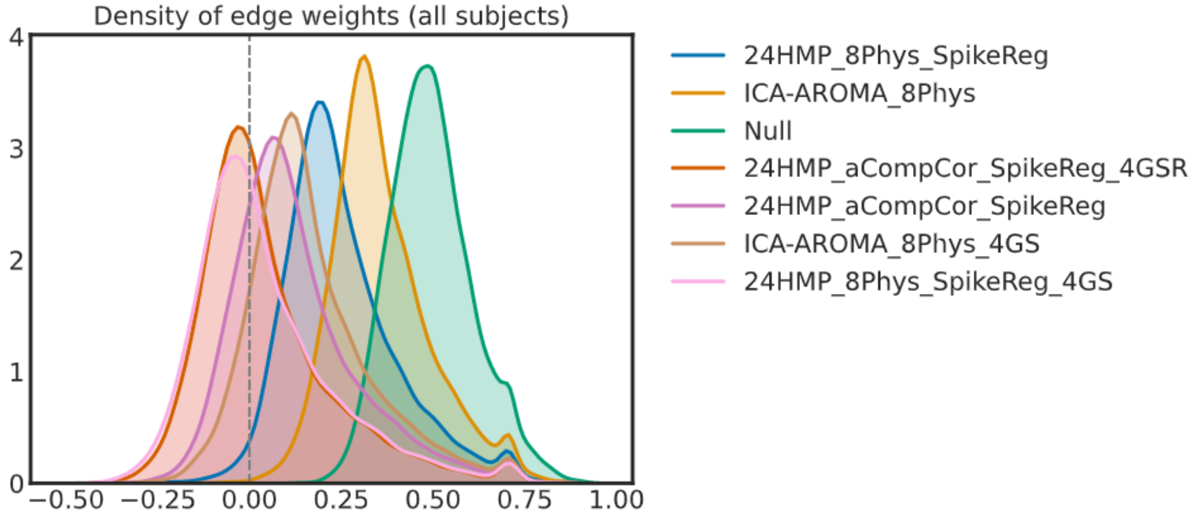


**Figure S2**. The density of edge weights across various denoising pipelines

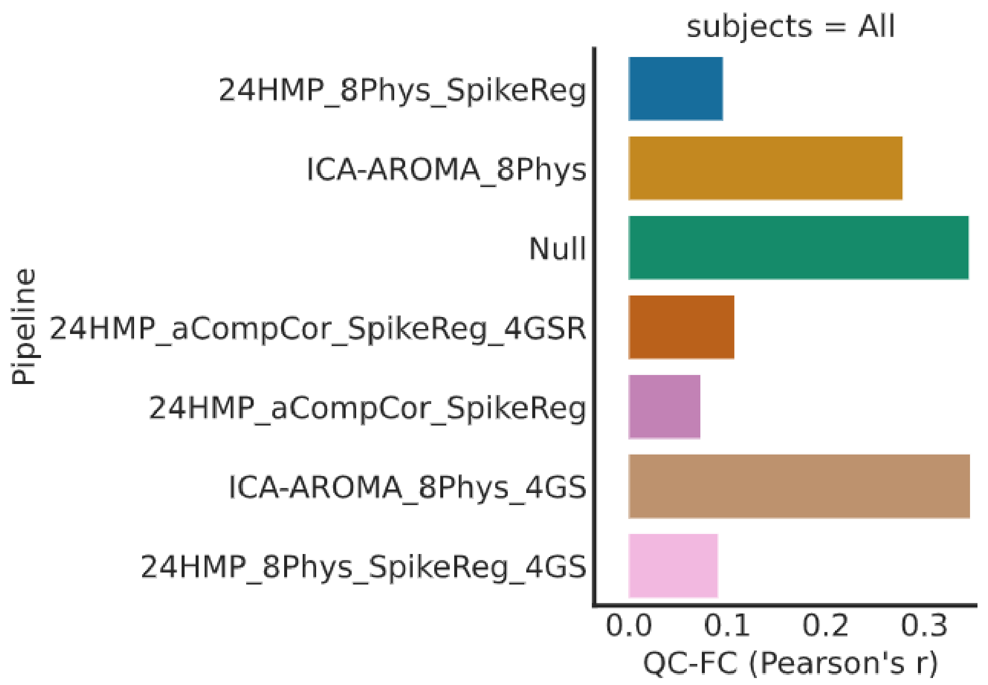


**Figure S3**. The average correlation between average movement (FD) and functional connectivity at each edge.

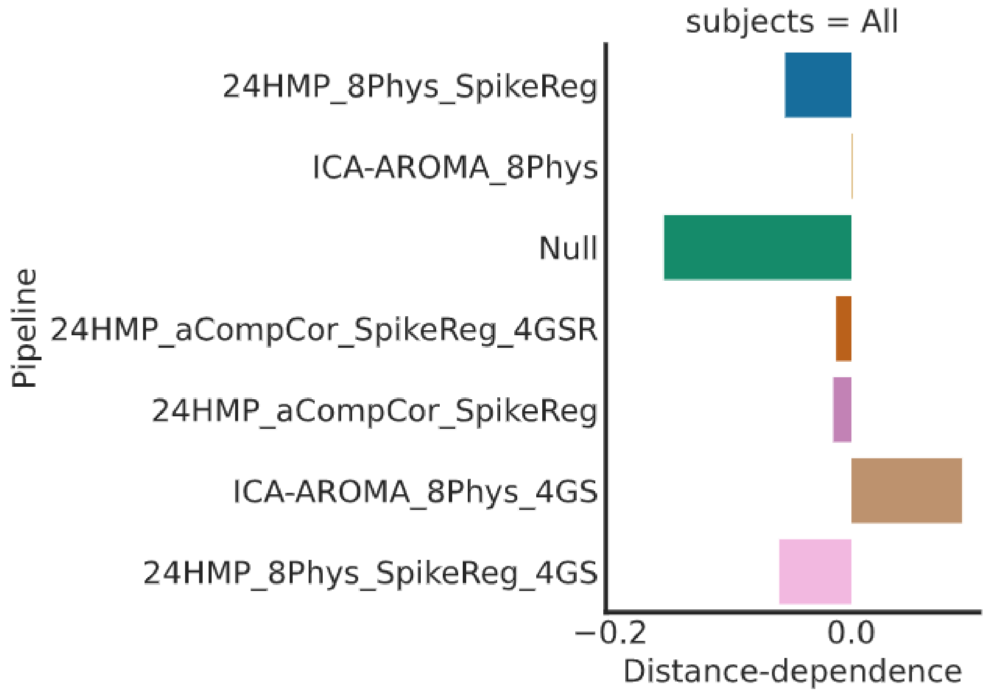


**Figure S4**. The average correlation between functional connectivity at each edge and the Euclidean distance between nodes.

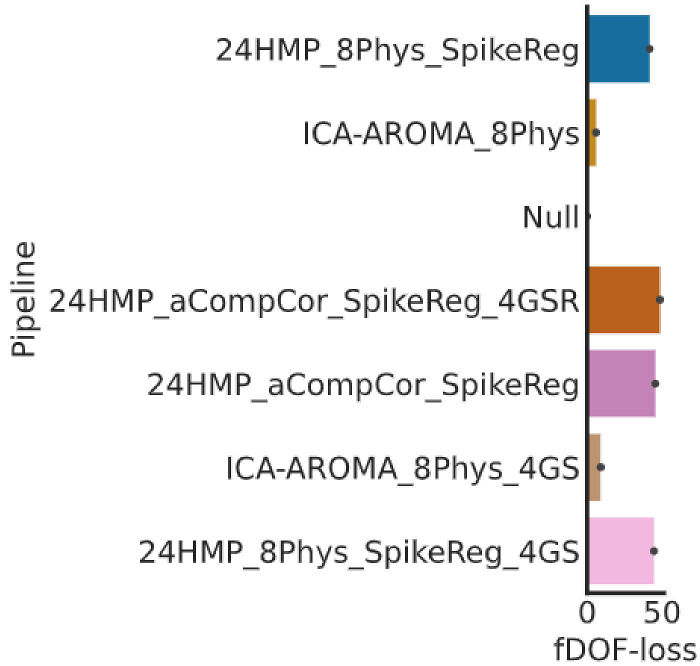


**Figure S5**. The functional degrees of freedom lost (fDOF) according to the number of confound regressors used.

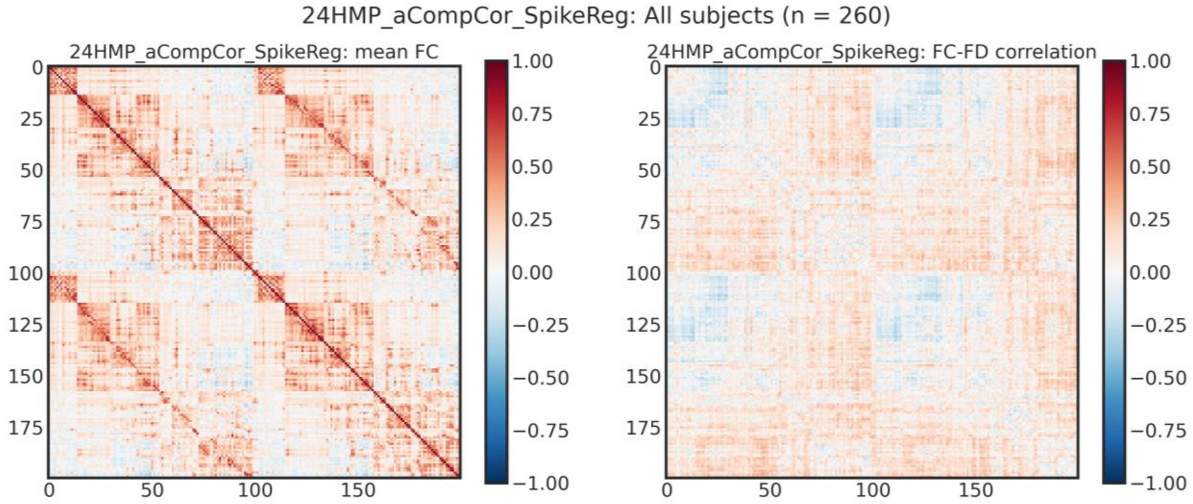


**Figure S6**. The correlation matrix (Pearson's R) after the selected confound regression strategy including: 24 head movement parameters, 10 principal components from WM and CSF, and motion spikes (left). The corresponding correlation between average movement and functional connectivity at each edge (right). Note, the 200 region parcellation from Schaefer et al. (2018) was used here.

### Supplementary Results

#### Table S2. Behavioural profiles in the full sample

| | Controls | | C1 | | C2 | | C3 | |
|---|---|---|---|---|---|---|---|---|
| Conners scale: | Median | MAD | Median | MAD | Median | MAD | Median | MAD |
| Inattention | 52.5 | 9.5 | 84 | 6 | 89 | 1 | 79 | 9.5 |
| Hyperactivity/Impulsivity | 53 | 8 | 88 | 2 | 81.5 | 8.5 | 59 | 11 |
| Learning Problems | 46 | 4 | 69 | 11 | 81 | 8 | 85 | 5 |
| Executive Function (cool) | 51 | 9 | 76 | 9 | 80.5 | 7.5 | 70 | 9 |
| Aggression | 46 | 2 | 84 | 6 | 52 | 7 | 54 | 9 |
| Peer Relations | 46 | 2 | 90 | 0 | 58 | 13 | 84 | 6 |

Note. Median scores and Median Absolute Deviation (MAD) on the Conners subscales for each group.

### Table S3. Pairwise group differences on the Conners scales in the full sample

| Conners scale: | Group 1 | Group 2 | Median Difference | *U* | *p* | Effect size |
|---|---|---|---|---|---|---|
| Inattention | Controls | C1 | 31.5 | 6672 | <0.001 | 0.875 |
| | Controls | C2 | 36.5 | 2742 | <0.001 | 0.933 |
| | Controls | C3 | 26.5 | 4704 | <0.001 | 0.833 |
| | C1 | C2 | 5 | 31730 | <0.001 | 0.64 |
| | C1 | C3 | -5 | 23987 | <0.001 | 0.602 |
| | C2 | C3 | -10 | 12081 | <0.001 | 0.739 |
| Hyperactivity/Impulsivity | Controls | C1 | 35 | 7522 | <0.001 | 0.86 |
| | Controls | C2 | 28.5 | 6918 | <0.001 | 0.832 |
| | Controls | C3 | 6 | 10948 | 0.001 | 0.611 |
| | C1 | C2 | -6.5 | 35883 | <0.001 | 0.593 |
| | C1 | C3 | -29 | 10676 | <0.001 | 0.823 |
| | C2 | C3 | -22.5 | 10529 | <0.001 | 0.772 |
| Learning Problems | Controls | C1 | 23 | 7424 | <0.001 | 0.861 |
| | Controls | C2 | 35 | 2020 | <0.001 | 0.951 |
| | Controls | C3 | 39 | 934 | <0.001 | 0.967 |
| | C1 | C2 | 12 | 26260 | <0.001 | 0.702 |
| | C1 | C3 | 16 | 13556 | <0.001 | 0.775 |
| | C2 | C3 | 4 | 18416 | <0.001 | 0.602 |
| Executive Function (cool) | Controls | C1 | 25 | 8577 | <0.001 | 0.84 |
| | Controls | C2 | 29.5 | 3289 | <0.001 | 0.92 |
| | Controls | C3 | 19 | 5704 | <0.001 | 0.797 |
| | C1 | C2 | 4.5 | 30395 | <0.001 | 0.655 |
| | C1 | C3 | -6 | 23790 | <0.001 | 0.606 |
| | C2 | C3 | -10.5 | 11082 | <0.001 | 0.761 |
| Aggression | Controls | C1 | 38 | 8880 | <0.001 | 0.834 |
| | Controls | C2 | 6 | 19366 | 0.961 | 0.529 |
| | Controls | C3 | 8 | 10976 | 0.001 | 0.61 |
| | C1 | C2 | -32 | 14856 | <0.001 | 0.831 |
| | C1 | C3 | -30 | 13620 | <0.001 | 0.774 |
| | C2 | C3 | 2 | 19098 | 0.005 | 0.587 |
| Peer Relations | Controls | C1 | 44 | 7768 | <0.001 | 0.855 |
| | Controls | C2 | 12 | 14327 | <0.001 | 0.651 |
| | Controls | C3 | 38 | 5310 | <0.001 | 0.811 |
| | C1 | C2 | -32 | 20302 | <0.001 | 0.77 |
| | C1 | C3 | -6 | 24754 | <0.001 | 0.59 |
| | C2 | C3 | 26 | 13763 | <0.001 | 0.703 |

Note. The table dispalys the pariwise median differences (Group2-Group1) between the groups on each subscale of the Conners. Mann-Whitney U-tests assess whether two samples come from different population distributions. The common language effect size is the probability that a randomly selected sample from one group has a higher score than a random sample from the other group. P-values are Bonferroni corrected for the total number of group comparisons.

**Table S4. Behavioural profiles in the MRI sample**

| | Controls | | C1 | | C2 | | C3 | |
|---|---|---|---|---|---|---|---|---|
| Conners scale: | Median | MAD | Median | MAD | Median | MAD | Median | MAD |
| Inattention | 50.5 | 10 | 84 | 6 | 90 | 0 | 81 | 9 |
| Hyperactivity/Impulsivity | 51 | 7 | 85.5 | 4.5 | 80 | 10 | 60 | 13 |
| Learning Problems | 46 | 4 | 67 | 11 | 81 | 9 | 88 | 2 |
| Executive Function (cool) | 46 | 6 | 73.5 | 9.5 | 80.5 | 6.5 | 72 | 8 |
| Aggression | 46 | 1 | 69.5 | 17.5 | 48.5 | 4.5 | 53 | 8 |
| Peer Relations | 46 | 1 | 69.5 | 17.5 | 48.5 | 4.5 | 53 | 8 |

Note. Median scores and Median Absolute Deviation (MAD) on the Conners subscales for each group.

### Table S5. Pairwise group differences on the Conners scales in the MRI sample

| Conners scale: | Group 1 | Group 2 | Median difference | *U* | *p* | Effect size |
|---|---|---|---|---|---|---|
| Inattention | Controls | C1 | 33.5 | 692 | <0.001 | 0.84 |
| | Controls | C2 | 39.5 | 270 | <0.001 | 0.922 |
| | Controls | C3 | 30.5 | 454 | <0.001 | 0.844 |
| | C1 | C2 | 6 | 1096 | <0.001 | 0.72 |
| | C1 | C3 | -3 | 1544 | 1 | 0.531 |
| | C2 | C3 | -9 | 704 | <0.001 | 0.733 |
| Hyperactivity/Impulsivity | Controls | C1 | 34.5 | 680 | <0.001 | 0.843 |
| | Controls | C2 | 29 | 599 | <0.001 | 0.827 |
| | Controls | C3 | 9 | 1061 | 0.046 | 0.636 |
| | C1 | C2 | -5.5 | 1746 | 0.85 | 0.555 |
| | C1 | C3 | -25.5 | 708 | <0.001 | 0.785 |
| | C2 | C3 | -20 | 660 | <0.001 | 0.749 |
| Learning Problems | Controls | C1 | 21 | 756 | <0.001 | 0.826 |
| | Controls | C2 | 35 | 232 | <0.001 | 0.933 |
| | Controls | C3 | 42 | 138 | <0.001 | 0.952 |
| | C1 | C2 | 14 | 1012 | <0.001 | 0.742 |
| | C1 | C3 | 21 | 500 | <0.001 | 0.848 |
| | C2 | C3 | 7 | 896 | 0.014 | 0.66 |
| Executive Function (cool) | Controls | C1 | 27.5 | 693 | <0.001 | 0.84 |
| | Controls | C2 | 34.5 | 265 | <0.001 | 0.924 |
| | Controls | C3 | 26 | 408 | <0.001 | 0.86 |
| | C1 | C2 | 7 | 1104 | <0.001 | 0.718 |
| | C1 | C3 | -1.5 | 1614 | 1 | 0.509 |
| | C2 | C3 | -8.5 | 760 | <0.001 | 0.711 |
| Aggression | Controls | C1 | 23.5 | 768 | <0.001 | 0.823 |
| | Controls | C2 | 2.5 | 1574 | 1 | 0.547 |
| | Controls | C3 | 7 | 1056 | 0.04 | 0.638 |
| | C1 | C2 | -21 | 818 | <0.001 | 0.791 |
| | C1 | C3 | -16.5 | 984 | <0.001 | 0.701 |
| | C2 | C3 | 4.5 | 1069 | 0.297 | 0.594 |
| Peer Relations | Controls | C1 | 43.5 | 594 | <0.001 | 0.863 |
| | Controls | C2 | 7 | 1384 | 1 | 0.601 |
| | Controls | C3 | 40 | 564 | <0.001 | 0.807 |
| | C1 | C2 | -36.5 | 986 | <0.001 | 0.749 |
| | C1 | C3 | -3.5 | 1454 | 0.798 | 0.558 |
| | C2 | C3 | 33 | 810 | 0.002 | 0.692 |

Note. The table dispalys the pariwise median differences (Group2-Group1) between the groups on each subscale of the Conners. Mann-Whitney U-tests assess whether two samples come from different population distributions. The common language effect size is the probability that a randomly selected sample from one group has a higher score than a random sample from the other group. P-values are Bonferroni corrected for the total number of group comparisons.

### Table S6. Pairwise chi-square tests of gender and diagnoses in the full sample

| | Group 1 | Group 2 | Percent difference | $X^2$ | *p* |
|---|---|---|---|---|---|
| Gender | Controls | C1 | 23.61 | 28.92 | <0.001 |
| | Controls | C2 | 3.29 | 0.31 | 1 |
| | Controls | C3 | 5.47 | 0.82 | 1 |
| | C1 | C2 | -20.33 | 28.61 | <0.001 |
| | C1 | C3 | -18.14 | 18.89 | <0.001 |
| | C2 | C3 | 2.18 | 0.13 | 1 |
| Diagnosis | Controls | C1 | 44.29 | 92.22 | <0.001 |
| | Controls | C2 | 28.13 | 44.73 | <0.001 |
| | Controls | C3 | 33.28 | 53.11 | <0.001 |
| | C1 | C2 | -16.16 | 15.22 | 0.001 |
| | C1 | C3 | -11 | 5.29 | 0.129 |
| | C2 | C3 | 5.16 | 1.03 | 1 |
| ADHD | Controls | C1 | 35.65 | 71.26 | <0.001 |
| | Controls | C2 | 17.06 | 26.98 | <0.001 |
| | Controls | C3 | 11.16 | 15.28 | 0.001 |
| | C1 | C2 | -18.59 | 24.2 | <0.001 |
| | C1 | C3 | -24.49 | 33.61 | <0.001 |
| | C2 | C3 | -5.89 | 2.4 | 0.729 |
| Dyslexia | Controls | C1 | 0.8 | 0.07 | 1 |
| | Controls | C2 | 6.43 | 6.9 | 0.052 |
| | Controls | C3 | 9.41 | 11.09 | 0.005 |
| | C1 | C2 | 5.63 | 9.56 | 0.012 |
| | C1 | C3 | 8.61 | 16.36 | <0.001 |
| | C2 | C3 | 2.98 | 0.82 | 1 |
| Autism | Controls | C1 | 10.62 | 16.54 | <0.001 |
| | Controls | C2 | 3.08 | 3.45 | 0.379 |
| | Controls | C3 | 7.3 | 10.12 | 0.009 |
| | C1 | C2 | -7.54 | 11.22 | 0.005 |
| | C1 | C3 | -3.32 | 1.13 | 1 |
| | C2 | C3 | 4.23 | 3.26 | 0.426 |

Note. The table displays group differences in percentages of boys and diagnoses (Group2-Group1), and the chi square test of difference between expected and observed frequencies. P-values are Bonferroni corrected for the total number of group comparisons.

**Table S7. Pairwise t-tests of age, IQ, maths and reading in the full sample**

| | Group 1 | Group 2 | Mean difference | *t* | *p* | Cohen's *d* |
|---|---|---|---|---|---|---|
| Age | Controls | C1 | -0.42 | 0.96 | 1 | 0.18 |
| | Controls | C2 | -0.11 | 0.28 | 1 | 0.05 |
| | Controls | C3 | -1.03 | 2.31 | 0.137 | 0.48 |
| | C1 | C2 | 0.31 | -0.79 | 1 | -0.14 |
| | C1 | C3 | -0.61 | 1.38 | 1 | 0.27 |
| | C2 | C3 | -0.92 | 2.26 | 0.154 | 0.44 |
| IQ | Controls | C1 | -13.06 | 11.05 | 0.001 | 1.09 |
| | Controls | C2 | -16.26 | 13.85 | 0.001 | 1.41 |
| | Controls | C3 | -20.72 | 15.24 | 0.001 | 1.68 |
| | C1 | C2 | -3.2 | 3.15 | 0.01 | 0.26 |
| | C1 | C3 | -7.65 | 6.4 | 0.001 | 0.59 |
| | C2 | C3 | -4.46 | 3.68 | 0.002 | 0.36 |
| Reading | Controls | C1 | -17.07 | 10.91 | <0.001 | 1.12 |
| | Controls | C2 | -22.17 | 15.16 | <0.001 | 1.57 |
| | Controls | C3 | -29.25 | 18.69 | <0.001 | 2.07 |
| | C1 | C2 | -5.1 | 3.69 | 0.001 | 0.31 |
| | C1 | C3 | -12.18 | 7.79 | <0.001 | 0.74 |
| | C2 | C3 | -7.08 | 4.72 | <0.001 | 0.46 |
| Maths | Controls | C1 | -25.85 | 14.76 | <0.001 | 1.43 |
| | Controls | C2 | -32.79 | 20.54 | <0.001 | 2.02 |
| | Controls | C3 | -35.94 | 20.18 | <0.001 | 2.21 |
| | C1 | C2 | -6.94 | 5.08 | <0.001 | 0.43 |
| | C1 | C3 | -10.09 | 6.4 | <0.001 | 0.62 |
| | C2 | C3 | -3.15 | 2.26 | 0.145 | 0.22 |
| WIAT-II Maths | Controls | C1 | -25.16 | 14.09 | <0.001 | 1.38 |
| | Controls | C2 | -32.38 | 19.65 | <0.001 | 1.98 |
| | Controls | C3 | -35.35 | 19.16 | <0.001 | 2.17 |
| | C1 | C2 | -7.22 | 4.99 | <0.001 | 0.44 |
| | C1 | C3 | -10.19 | 6.08 | <0.001 | 0.62 |
| | C2 | C3 | -2.98 | 2.01 | 0.271 | 0.21 |

Note. The table displays the pariwise mean differences (Group2-Group1) between the groups. Maths scores are reported for a subset of children who completed the Numeric Operations subtest of Wechsler Individual Achievement Test II (Maths WIAT-II) and for the whole sample using a combined normalized maths score (Maths), which combines z-scored Numeric Operations WIAT-II with z-scored Maths Fluency subtest of Woodcock Johnson III Test of Achievement (WJ-III). P-values are Bonferroni corrected for the total number of group comparisons.

**Table S8. Pairwise chi-square tests of gender and diagnoses in the MRI sample**

| | Group 1 | Group 2 | Percent difference | Chi2 | p |
|---|---|---|---|---|---|
| Gender | Controls | C1 | 33.41 | 14.32 | 0.001 |
| | Controls | C2 | 13.77 | 1.72 | 1 |
| | Controls | C3 | 10.16 | 0.73 | 1 |
| | C1 | C2 | -19.64 | 4.8 | 0.17 |
| | C1 | C3 | -23.25 | 6.09 | 0.082 |
| | C2 | C3 | -3.61 | 0.03 | 1 |
| Diagnosis | Controls | C1 | 36.59 | 22.02 | <0.001 |
| | Controls | C2 | 25.52 | 11.86 | 0.003 |
| | Controls | C3 | 35.59 | 18.87 | <0.001 |
| | C1 | C2 | -11.07 | 1.2 | 1 |
| | C1 | C3 | -1 | 0.01 | 1 |
| | C2 | C3 | 10.07 | 0.74 | 1 |
| ADHD | Controls | C1 | 29.82 | 18.29 | <0.001 |
| | Controls | C2 | 12.67 | 5.03 | 0.15 |
| | Controls | C3 | 6.9 | 1.54 | 1 |
| | C1 | C2 | -17.14 | 4.14 | 0.251 |
| | C1 | C3 | -22.92 | 7.27 | 0.042 |
| | C2 | C3 | -5.78 | 0.36 | 1 |
| Dyslexia | Controls | C1 | 1.06 | 0.02 | 1 |
| | Controls | C2 | 11.06 | 3.32 | 0.41 |
| | Controls | C3 | 9.54 | 2.31 | 0.77 |
| | C1 | C2 | 10 | 2.75 | 0.583 |
| | C1 | C3 | 8.48 | 1.78 | 1 |
| | C2 | C3 | -1.52 | 0 | 1 |
| Autism | Controls | C1 | 11.43 | 5.67 | 0.104 |
| | Controls | C2 | 1.79 | 0 | 1 |
| | Controls | C3 | 8.51 | 3.33 | 0.407 |
| | C1 | C2 | -9.64 | 3.03 | 0.491 |
| | C1 | C3 | -2.92 | 0.04 | 1 |
| | C2 | C3 | 6.72 | 1.26 | 1 |

Note. The table displays group differences in percentages of boys and diagnoses (Group2-Group1), and the chi square test of difference between expected and observed frequencies P-values are Bonferroni corrected for the total number of group comparisons.

### Table S9. Pairwise t-tests of group differences in the MRI sample

| | Group 1 | Group 2 | Mean difference | t | p | Cohen's *d* |
|---|---|---|---|---|---|---|
| Age | Controls | C1 | -0.06 | 0.16 | 1 | 0.03 |
| | Controls | C2 | -0.72 | 2.02 | 0.275 | 0.37 |
| | Controls | C3 | -0.51 | 1.21 | 1 | 0.23 |
| | C1 | C2 | -0.65 | 1.66 | 0.6 | 0.3 |
| | C1 | C3 | -0.45 | 0.98 | 1 | 0.19 |
| | C2 | C3 | 0.21 | -0.51 | 1 | -0.1 |
| IQ | Controls | C1 | -9.96 | 4.86 | <0.001 | 0.85 |
| | Controls | C2 | -15.9 | 7.94 | <0.001 | 1.46 |
| | Controls | C3 | -20.92 | 7.78 | <0.001 | 1.46 |
| | C1 | C2 | -5.94 | 2.75 | 0.042 | 0.5 |
| | C1 | C3 | -10.96 | 3.94 | 0.001 | 0.72 |
| | C2 | C3 | -5.03 | 1.77 | 0.475 | 0.34 |
| Reading | Controls | C1 | -13.32 | 4.98 | <0.001 | 0.89 |
| | Controls | C2 | -22.13 | 8.78 | <0.001 | 1.62 |
| | Controls | C3 | -30.03 | 11.24 | <0.001 | 2.15 |
| | C1 | C2 | -8.81 | 2.92 | 0.025 | 0.53 |
| | C1 | C3 | -16.71 | 5.14 | <0.001 | 0.99 |
| | C2 | C3 | -7.9 | 2.53 | 0.077 | 0.5 |
| Maths | Controls | C1 | -21.47 | 5.93 | <0.001 | 1.04 |
| | Controls | C2 | -33.2 | 10.38 | <0.001 | 1.93 |
| | Controls | C3 | -36.38 | 10.27 | <0.001 | 2.04 |
| | C1 | C2 | -11.72 | 3.63 | 0.002 | 0.67 |
| | C1 | C3 | -14.91 | 4.18 | <0.001 | 0.82 |
| | C2 | C3 | -3.19 | 1.14 | 1 | 0.23 |
| WIAT-II Maths | Controls | C1 | -20.66 | 5.64 | <0.001 | 1 |
| | Controls | C2 | -31.83 | 9.26 | <0.001 | 1.84 |
| | Controls | C3 | -33.66 | 8.81 | <0.001 | 1.9 |
| | C1 | C2 | -11.17 | 3.19 | 0.011 | 0.63 |
| | C1 | C3 | -13 | 3.34 | 0.007 | 0.72 |
| | C2 | C3 | -1.83 | 0.59 | 1 | 0.13 |
| Mean FC | Controls | C1 | 0 | -0.48 | 1 | -0.08 |
| | Controls | C2 | 0.01 | -2.26 | 0.154 | -0.42 |
| | Controls | C3 | 0 | 0.27 | 1 | 0.05 |
| | C1 | C2 | 0.01 | -1.73 | 0.518 | -0.31 |
| | C1 | C3 | 0 | 0.67 | 1 | 0.13 |
| | C2 | C3 | -0.01 | 2.22 | 0.173 | 0.44 |
| Framewise displacement | Controls | C1 | 0.06 | -3.82 | 0.001 | -0.67 |
| | Controls | C2 | 0.04 | -2.81 | 0.035 | -0.52 |
| | Controls | C3 | 0.04 | -2.59 | 0.066 | -0.49 |
| | C1 | C2 | -0.02 | 0.99 | 1 | 0.18 |
| | C1 | C3 | -0.02 | 1.04 | 1 | 0.2 |
| | C2 | C3 | 0 | 0.11 | 1 | 0.02 |

Note. The table displays the pariwise mean differences (Group2-Group1) between the groups. Maths scores are reported for a subset of children who completed the Numeric Operations subtest of Wechsler Individual Achievement Test II (Maths WIAT-II) and for the whole sample using a combined normalized maths score (Maths), which combines z-scored Numeric Operations WIAT-II with z-scored Maths Fluency subtest of Woodcock Johnson III Test of Achievement (WJ-III). P-values are Bonferroni corrected for the total number of group comparisons.

#### Table S10. Group effects on global graph metrics across all group thresholds

| | F-ratio AUC | Permuted AUC: M (SD) | *p* |
|---|---|---|---|
| Strength | 6.10 | 5.47 (8.53) | 0.291 |
| Modularity | 9.65 | 5.31 (6.1) | 0.167 |
| Path length | 8.53 | 5.23 (4.38) | 0.171 |
| Global efficiency | 12.23 | 5.07 (6.93) | 0.103 |
| Local efficiency | 3.79 | 5.32 (6.65) | 0.429 |
| Clustering | 6.49 | 5.37 (7.02) | 0.263 |
| Small-worldness | 4.97 | 5.19 (5.09) | 0.367 |
| Assortativity | 0.4 | 4.87 (6.31) | 0.903 |

*Note.* All graph metrics were normalised according to 100 random graphs except for strength and modularity.

#### Table S11. Group effects on global graph metrics across all individual thresholds

| | F-ratio AUC | Permuted AUC: M (SD) | *p* |
|---|---|---|---|
| Strength | 0.23 | 5.22 (7.71) | 0.852 |
| Modularity | 9.72 | 5.55 (7.34) | 0.172 |
| Path length | 0.65 | 5.11 (6.62) | 0.815 |
| Global efficiency | 0.2 | 5.05 (6.77) | 0.943 |
| Local efficiency | 2.44 | 4.81 (5.99) | 0.534 |
| Clustering | 3.91 | 4.79 (5.68) | 0.397 |
| Small-worldness | 3.59 | 4.75 (5.62) | 0.42 |
| Assortativity | 0.99 | 4.74 (6) | 0.682 |

*Note.* All graph metrics were normalised according to 100 random graphs except for strength and modularity.

**Table S12. PLS of ICN Connectivity across connectome thresholds**

| | | ICN Connectivity | | Permutation Test | |
|---|---|---|---|---|---|
| Threshold (%) | PLS Components | RMSE (SEM) | Accuracy (SEM) | RMSE (SEM) | *p* |
| 5 | 1 | 0.481 (0.017) | 0.32 (0.024) | 0.512 (0.018) | 0.055 |
| 10 | 2 | 0.475 (0.015) | 0.328 (0.022) | 0.517 (0.019) | 0.016 |
| 15 | 3 | 0.461 (0.015) | 0.348 (0.022) | 0.52 (0.02) | 0.002 |
| 20 | 3 | 0.482 (0.02) | 0.318 (0.028) | 0.52 (0.02) | 0.036 |
| 25 | 3 | 0.471 (0.011) | 0.334 (0.015) | 0.52 (0.02) | 0.01 |
| 30 | 4 | 0.465 (0.015) | 0.342 (0.022) | 0.522 (0.019) | 0.002 |

**Table S13. PLS of Nodal Strength across connectome thresholds**

| | | Nodal Strength | | Permutation Test | |
|---|---|---|---|---|---|
| Threshold (%) | PLS Components | RMSE (SEM) | Accuracy (SEM) | RMSE (SEM) | *p* |
| 5 | 3 | 0.475 (0.016) | 0.328 (0.022) | 0.518 (0.018) | 0.014 |
| 10 | 7 | 0.471 (0.017) | 0.334 (0.024) | 0.522 (0.018) | 0.002 |
| 15 | 1 | 0.488 (0.009) | 0.31 (0.013) | 0.513 (0.019) | 0.112 |
| 20 | 6 | 0.471 (0.013) | 0.334 (0.019) | 0.52 (0.018) | 0.005 |
| 25 | 6 | 0.474 (0.014) | 0.33 (0.02) | 0.52 (0.018) | 0.007 |
| 30 | 1 | 0.49 (0.016) | 0.306 (0.022) | 0.512 (0.018) | 0.128 |

**Table S14. PLS of Connector Hub Strength across connectome thresholds**

| | | | Connector Hub Strength | | Permutation Test | |
|---|---|---|---|---|---|---|
| Threshold (%) | Hubs | PLS Components | RMSE (SEM) | Accuracy (SEM) | RMSE (SEM) | *p* |
| 5 | 36 | 1 | 0.488 (0.011) | 0.31 (0.015) | 0.511 (0.017) | 0.108 |
| 10 | 39 | 1 | 0.469 (0.013) | 0.336 (0.018) | 0.51 (0.018) | 0.017 |
| 15 | 34 | 2 | 0.459 (0.015) | 0.351 (0.021) | 0.515 (0.018) | <0.001 |
| 20 | 30 | 2 | 0.476 (0.017) | 0.327 (0.024) | 0.515 (0.018) | 0.022 |
| 25 | 31 | 1 | 0.463 (0.016) | 0.346 (0.023) | 0.515 (0.018) | 0.003 |
| 30 | 31 | 1 | 0.483 (0.016) | 0.316 (0.023) | 0.509 (0.017) | 0.073 |

**Table S15. PLS of Provinicial Hub Strength across connectome thresholds**

| | | | Provincial Hub Strength | | Permutation Test | |
|---|---|---|---|---|---|---|
| Threshold (%) | Hubs | PLS Components | RMSE (SEM) | Accuracy (SEM) | RMSE (SEM) | *p* |
| 5 | 122 | 4 | 0.467 (0.019) | 0.339 (0.027) | 0.519 (0.018) | 0.006 |
| 10 | 114 | 4 | 0.471 (0.015) | 0.333 (0.021) | 0.518 (0.018) | 0.008 |
| 15 | 106 | 6 | 0.484 (0.016) | 0.315 (0.022) | 0.52 (0.019) | 0.033 |
| 20 | 104 | 7 | 0.465 (0.02) | 0.343 (0.028) | 0.521 (0.019) | 0.003 |
| 25 | 103 | 7 | 0.468 (0.017) | 0.338 (0.024) | 0.521 (0.019) | 0.004 |
| 30 | 104 | 6 | 0.49 (0.018) | 0.306 (0.025) | 0.52 (0.019) | 0.069 |

### Additional Components of the PLS

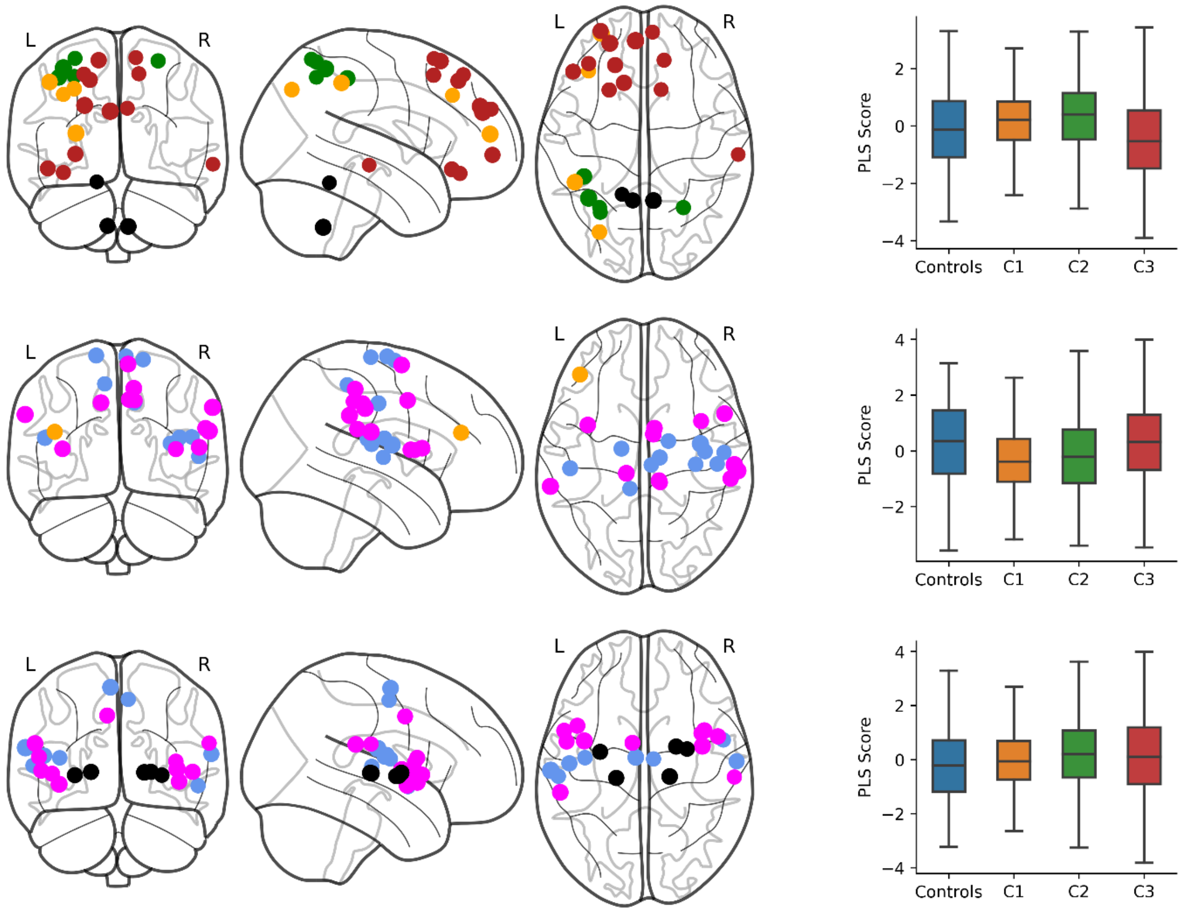


**Figure S7.** PLS components 3-5 of nodal strength (top to bottom) that predicted group membership. The brain plots (left) show the 5% largest loadings of nodes onto each component relative to their standard error over 1000 bootstrapped samples. The size of the node is proportional to its absolute loading and the colour corresponds to its ICN: default-mode (red), fronto-parietal (orange), dorsal attention (green), subcortical (black), ventral attention (pink), and somatomotor (blue). The boxplots (right) show the bootstrapped distribution of component scores for each group and significant group differences assessed by a permutation test.

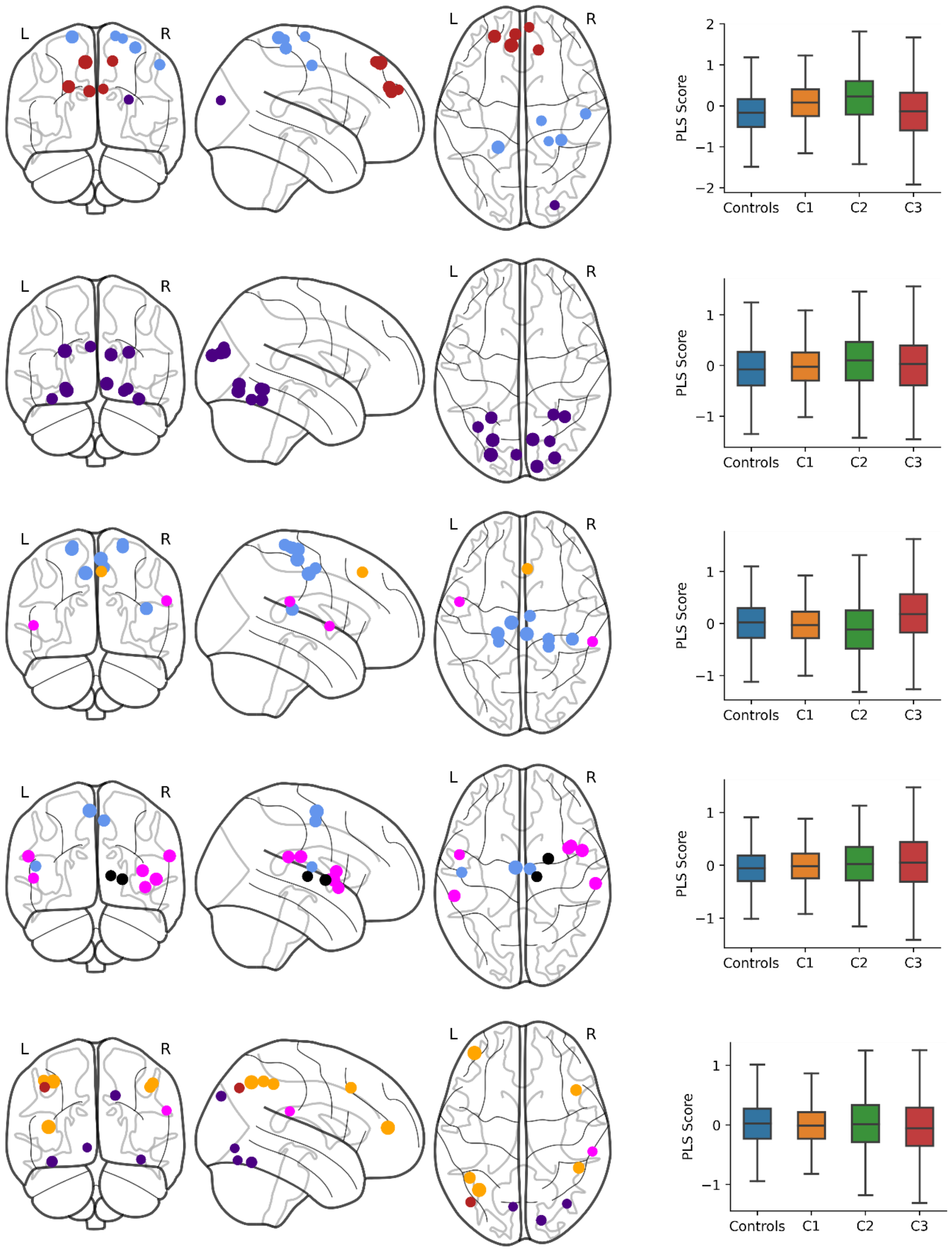


**Figure S8.** PLS components 3-7 of provincial hub strength (top to bottom) that predicted group membership. The brain plots (left) show the 10% largest loadings of nodes onto each component relative to their standard error over 1000 bootstrapped samples. The size of the node is proportional to its absolute loading and the colour corresponds to its ICN: default-mode (red), somatomotor (blue), visual (purple), ventral attention (pink), fronto-parietal (orange), and subcortical (black). The boxplots (right) show the bootstrapped distribution of component scores for each group and significant group differences assessed by a permutation test.
